# Machine Learning for Identifying Data-Driven Subphenotypes of Incident Post-Acute SARS-CoV-2 Infection Conditions with Large Scale Electronic Health Records: Findings from the RECOVER Initiative

**DOI:** 10.1101/2022.05.21.22275412

**Authors:** Hao Zhang, Chengxi Zang, Zhenxing Xu, Yongkang Zhang, Jie Xu, Jiang Bian, Dmitry Morozyuk, Dhruv Khullar, Yiye Zhang, Anna S. Nordvig, Edward J. Schenck, Elizabeth A. Shenkman, Russel L. Rothman, Jason P. Block, Kristin Lyman, Mark G. Weiner, Thomas W. Carton, Fei Wang, Rainu Kaushal

## Abstract

The post-acute sequelae of SARS-CoV-2 infection (PASC) refers to a broad spectrum of symptoms and signs that are persistent, exacerbated, or newly incident in the post-acute SARS-CoV-2 infection period of COVID-19 patients. Most studies have examined these conditions individually without providing concluding evidence on co-occurring conditions. To answer this question, this study leveraged electronic health records (EHRs) from two large clinical research networks from the national Patient-Centered Clinical Research Network (PCORnet) and investigated patients’ newly incident diagnoses that appeared within 30 to 180 days after a documented SARS-CoV-2 infection. Through machine learning, we identified four reproducible subphenotypes of PASC dominated by blood and circulatory system, respiratory, musculoskeletal and nervous system, and digestive system problems, respectively. We also demonstrated that these subphenotypes were associated with distinct patterns of patient demographics, underlying conditions present prior to SARS-CoV-2 infection, acute infection phase severity, and use of new medications in the post-acute period. Our study provides novel insights into the heterogeneity of PASC and can inform stratified decision-making in the treatment of COVID-19 patients with PASC conditions.

## Introduction

A variety of symptoms and signs involving multiple organ systems (e.g., cardiovascular^1^, mental^2^, metabolic^3^, and renal^4^) were persistent, exacerbated or newly developed following the acute phase of SARS-CoV-2 infection. Currently, our understanding of these conditions, typically regarded as post-acute sequelae of SARS-CoV-2 infection (PASC)^5,6^, remains limited. Most existing studies have investigated these PASC conditions individually (e.g., by examining the incidence^7^ or excess burden^8^ of each symptom or condition in the post-acute period for COVID-19 patients relative to controls). It is unclear if any of these PASC symptoms and conditions tend to co-appear together or are more likely to develop in certain patient populations.

To answer the above question, we developed a machine learning approach to derive subphenotypes of SARS-CoV-2 infected patients based on the newly incident conditions in the post-acute period (defined as 30 to 180 days after their confirmed SARS-CoV-2 infection) using the data from electronic health records (EHR). We focused on new incidences in this study because it provided a clean way of defining PASC phenotypes without complicated consideration of pre-existing conditions. We leveraged the EHR repositories from two large-scale clinical research networks (CRNs) from the national Patient-Centered Clinical Research Network (PCORnet): the INSIGHT network^9^, which includes 12 million patients in the New York City (NYC) area, and the OneFlorida+ network^10^, which includes 19 million patients from Florida, Georgia, and Alabama. We examined the incidence of 137 diagnosis categories defined from the Clinical Classifications Software Refined (CCSR) categories^11^. Four distinct subphenotypes were identified from the INSIGHT CRN data and validated in the OneFlorida+ CRN data.

Patients in Subphenotype 1 were older with incident blood and circulatory conditions in the post-acute phase. Subphenotype 2 included patients who are younger with incident respiratory problems. Subphenotype 3 included patients who developed musculoskeletal and nervous system conditions. Lastly, patients in Subphenotype 4 are characterized by digestive systems incident conditions. Our study dissects the heterogeneity of potential PASC conditions as different subgroups, which can inform the classification and treatment of PASC. This study is part of the NIH Researching COVID to Enhance Recovery (RECOVER) Initiative^12^, which seeks to understand, treat, and prevent the post-acute sequelae of SARS-CoV-2 infection (PASC). For more information on RECOVER, visit https://recovercovid.org/

## Results

### Overall Pipeline

Our overall analytics pipeline is demonstrated in Figure 1. Using the two CRNs, we extracted patients who had positive nucleic acid amplification or antigen viral tests for SARS-CoV-2 from March 2020 to November 2021. A list of 137 potential PASC diagnosis categories (Methods) were compiled and only patients who had documented new incidences of these conditions in their post-acute infection period were retained. Each patient was initially represented as a 137-dimensional binary vector according to whether a particular condition appeared in the post-acute infection period or not (Step 1). Then a set of “PASC topics” was learned based on the co-incidence patterns of these conditions (Step 2) and the initial patient vectors were projected onto these learned topics to obtain their topic loading representations (Step 3), which was further used in a clustering procedure to identify the subphenotypes (Step 4). Details on the concepts and methods involved in our pipeline are presented in Methods. In the following text, we present our main results.

**Figure 1.**
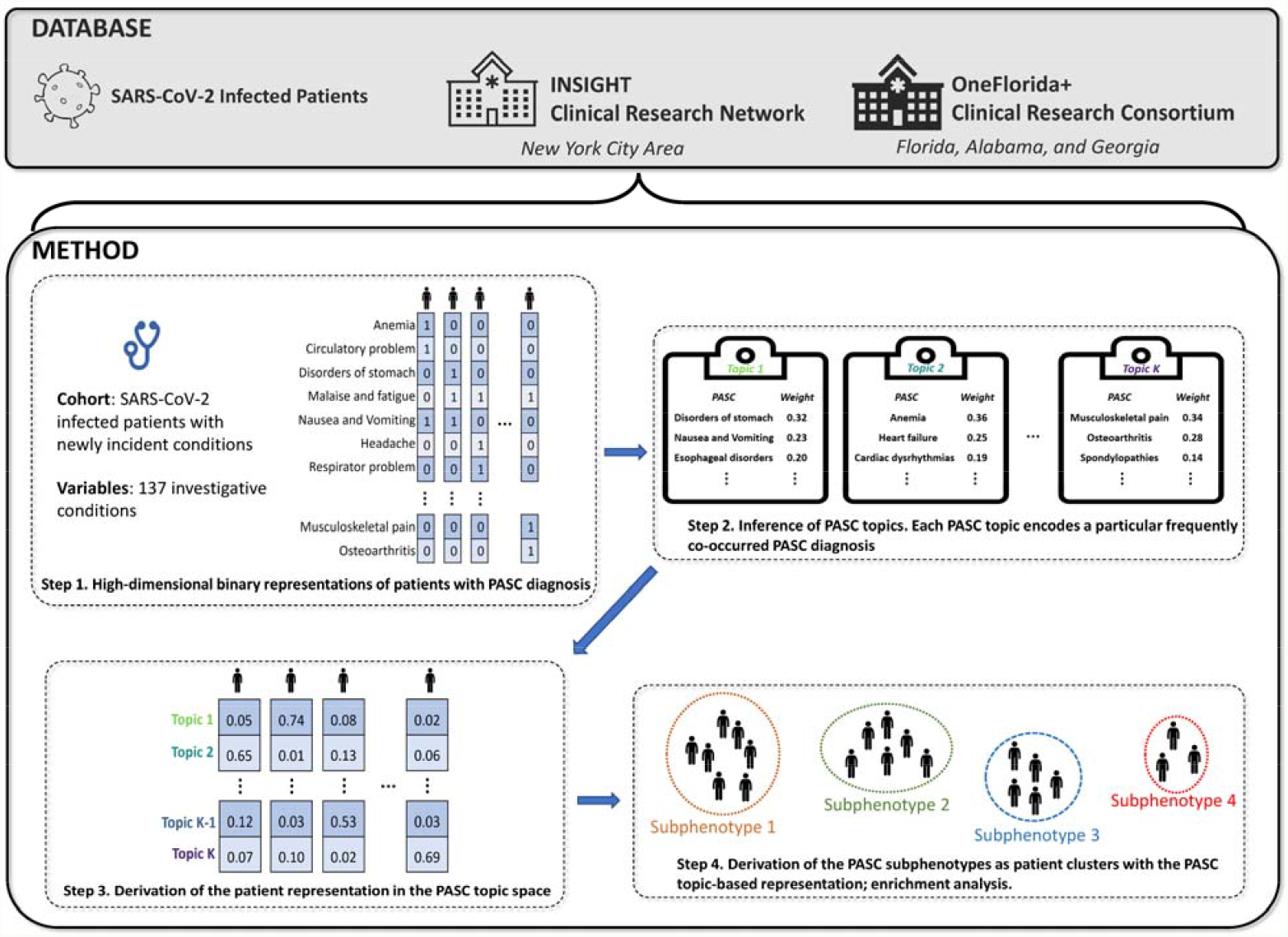
Data creation and subphenotype pipeline. To study the subphenotypes for patients with PASC, we constructed cohorts from the INSIGHT and OneFlorida+ clinical research networks. After obtaining high-dimensional binary representations of patients with PASC diagnoses (**step 1**), we learned PASC topics (**step 2**) and inferred the patient representations in the low-dimensional PASC topic space (**step 3**) by a topic modeling approach. Finally, we derived PASC subphenotypes as patient clusters with the PASC topic-based representations (**step 4**).

### Study Cohorts

Our study included 20,881 patients from the INSIGHT CRN and 13,724 patients from the OneFlorida+ CRN who tested positive for SARS-CoV-2 on viral tests (see Methods for detailed inclusion-exclusion criteria). The patients within the INSIGHT cohort had a median age of 58.0 (interquartile range [IQR] [42.0-70.0]) and a median Area Deprivation Index (ADI)^13^ of 15.0 (IQR [6.0-25.0]), consisting of 12,188 (58.37%) females, 7013 (33.59%) White patients, and 4771 (22.85%) Black patients. The OneFlorida+ cohort contained patients who were younger (median age of 51.0 (IQR [35.0-65.0])), with more disadvantaged social conditions (median ADI 59.0; IQR [42.0-76.0])), and more white patients (7175; 52.28%). 33.04% of the INSIGHT CRN patients had a confirmed SARS-CoV-2 infection from March to June 2020 (compared to 8.83% of the patients from OneFlorida+). This coincided with the first wave of COVID-19 in the US when NYC was the epicenter. Patients from OneFlorida+ were more likely to test positive from July to October 2020 (26% vs. 6% of patients from INSIGHT). Table 1 summarizes the summary statistics of the patients from the two cohorts.

**Table 1.** Characteristics of the INSIGHT cohort (for development) and the OneFlorida+ cohort (for validation).

| Characteristics | INSIGHT cohort<br>development | OneFlorida+ cohort<br>validation |
| --- | --- | --- |
| **No. of patients** | 20881 | 13724 |
| **Age, y, Median (IQR)<sup>c</sup>** | 58.0 [42.0-70.0] | 51.0 [35.0-65.0] |
| **Age group – no. (%)** |  |  |
| 20-<40 years | 4603 (22.04%) | 4284 (31.22%) |
| 40-<55 years | 4522 (21.66%) | 3318 (24.18%) |
| 55-<65 years | 4308 (20.63%) | 2528 (18.42%) |
| 65-<75 years | 3810 (18.25%) | 1923 (14.01%) |
| 75-<85 years | 2553 (12.22%) | 1178 (8.58%) |
| 85+ years | 1085 (5.20%) | 493 (3.59%) |
| **Sex – no. (%)** |  |  |
| Female | 12188 (58.37%) | 8468 (61.70%) |
| Male | 8692 (41.63%) | 5255 (38.29%) |
| Other/Missing | 1 (0.00%) | 1 (0.00%) |
| **Race – no. (%)** |  |  |
| Asian | 945 (4.52%) | 144 (1.05%) |
| Black or African American | 4771 (22.85%) | 4076 (29.70%) |
| White | 7013 (33.59%) | 7175 (52.28%) |
| Other | 6238 (29.87%) | 2123 (15.47%) |
| Missing | 1914 (9.17%) | 206 (1.50%) |
| **Ethnic group – no. (%)** |  |  |
| Hispanic: Yes | 6838 (32.74%) | 2881 (20.99%) |
| Hispanic: No | 12248 (58.66%) | 9329 (67.98%) |
| Hispanic: Other/Missing | 1795 (8.60%) | 1514 (11.03%) |
| **Area deprivation index, Median (IQR)** | 15.0 [6.0-25.0] | 59.0 [42.0-76.0] |
| **Healthcare utilization in the past 3 yr – no. (%)** |  |  |
| Inpatient 0 | 14631 (70.07%) | 7437 (54.19%) |
| Inpatient 1-2 | 4359 (20.88%) | 3018 (21.99%) |
| Inpatient 3-4 | 1074 (5.14%) | 1195 (8.71%) |
| Inpatient >=5 | 817 (3.91%) | 2074 (15.11%) |
| Outpatient 0 | 1241 (5.94%) | 2759 (20.10%) |
| Outpatient 1-2 | 2012 (9.64%) | 1736 (12.65%) |
| Outpatient 3-4 | 1595 (7.64%) | 820 (5.97%) |
| Outpatient >=5 | 16033 (76.78%) | 8409 (61.27%) |
| Emergency 0 | 11321 (54.22%) | 4908 (35.76%) |
| Emergency 1-2 | 5975 (28.61%) | 3386 (24.67%) |
| Emergency 3-4 | 1730 (8.29%) | 1465 (10.67%) |
| Emergency >=5 | 1855 (8.89%) | 3965 (28.89%) |
| **Index time period of patient – no. (%)** |  |  |
| 03/20-06/20 | 6899 (33.04%) | 1212 (8.83%) |
| 07/20-10/20 | 1180 (5.65%) | 3570 (26.01%) |
| 11/20-02/21 | 8714 (41.73%) | 3988 (29.06%) |
| 03/21-06/21 | 3390 (16.24%) | 1531 (11.16%) |
| 07/21-11/21 | 698 (3.34%) | 3423 (24.94%) |
| **Acute phase severities of COVID-19 (-1~16 days)<sup>a</sup> – no. (%)** |  |  |
| Hospitalized | 9076 (43.47%) | 5036 (36.69%) |
| Ventilation | 495 (2.37%) | 465 (3.39%) |
| Critical care | 1144 (5.48%) | 833 (6.07%) |
| **Underlying conditions<sup>b</sup> – no. (%)** |  |  |
| Alcohol Abuse | 697 (3.34%) | 503 (3.66%) |
| Anemia | 3288 (15.75%) | 2674 (19.48%) |
| Arrhythmia | 3656 (17.51%) | 2106 (15.35%) |
| Asthma | 2591 (12.41%) | 1641 (11.96%) |
| Cancer | 2406 (11.52%) | 1194 (8.70%) |
| Chronic Kidney Disease | 3223 (15.44%) | 2175 (15.85%) |
| Chronic Pulmonary Disorders | 3893 (18.64%) | 2778 (20.24%) |
| Cirrhosis | 408 (1.95%) | 241 (1.76%) |
| Coagulopathy | 1753 (8.39%) | 776 (5.65%) |
| Congestive Heart Failure | 2508 (12.01%) | 2003 (14.59%) |
| COPD | 1246 (5.97%) | 1125 (8.20%) |
| Coronary Artery Disease | 3234 (15.49%) | 1924 (14.02%) |
| Dementia | 1046 (5.01%) | 688 (5.01%) |
| Diabetes Type 1 | 287 (1.37%) | 265 (1.93%) |
| Diabetes Type 2 | 5173 (24.77%) | 3544 (25.82%) |
| End Stage Renal Disease on Dialysis | 1080 (5.17%) | 622 (4.53%) |
| Hemiplegia | 311 (1.49%) | 242 (1.76%) |
| HIV<sup>d</sup> | 376 (1.81%) | 123 (0.90%) |
| Hypertension | 9165 (43.89%) | 6486 (47.26%) |
| Hypertension and Type 1 or 2 Diabetes | 4367 (20.91%) | 3064 (22.33%) |
| Inflammatory Bowel Disorder | 215 (1.03%) | 145 (1.06%) |
| Lupus or SLE<sup>e</sup> | 186 (0.89%) | 173 (1.26%) |
| Mental Health Disorders | 2556 (12.24%) | 2559 (18.65%) |
| Multiple Sclerosis | 114 (0.55%) | 73 (0.53%) |
| Parkinson's Disease | 144 (0.68%) | 107 (0.78%) |
| Peripheral vascular disorders | 1791 (8.57%) | 1174 (8.55%) |
| Pregnant | 686 (3.29%) | 788 (5.74%) |
| Pulmonary Circulation Disorder | 467 (2.24%) | 342 (2.49%) |
| Rheumatoid Arthritis | 375 (1.79%) | 296 (2.16%) |
| Seizure/Epilepsy | 546 (2.61%) | 511 (3.72%) |
| Severe Obesity (BMI>=40 kg/m2) | 1600 (7.66%) | 1789 (13.04%) |
| Weight Loss | 1145 (5.48%) | 783 (5.71%) |
| Down's Syndrome | 20 (0.09%) | 20 (0.15%) |
| Other Substance Abuse | 1503 (7.20%) | 1857 (13.53%) |
| Cystic Fibrosis | 8 (0.04%) | 16 (0.12%) |
| Autism | 25 (0.12%) | 46 (0.34%) |
| Sickle Cell | 158 (0.76%) | 139 (1.01%) |
| Corticosteroids Prescription | 3193 (15.29%) | 2618 (19.08%) |
| Immunosuppressant Prescription | 1018 (4.88%) | 353 (2.57%) |

**Note:** a. Time range for acute phase severities is from one day before index date to sixteen days after index date; b. Coexisting conditions existed if two records in the 3-years prior to index event; c. IQR: inter-quartile range; d. HIV: human immunodeficiency virus; e. SLE: systemic lupus erythematosus; f. For the healthcare utilization in the past three years, including inpatient, outpatient, and emergency visits, we binned the number of visits into five levels.

### Potential PASC Topics

A list of 137 potentially PASC-related diagnoses groups defined by ICD-10 diagnosis codes and CCSR categories^11^ (Supplementary Table 1) was compiled for our study. We first investigated the co-incidence patterns across different diagnoses within 30-180 days after the SARS-CoV-2 infection confirmation for COVID-19 positive patients (Methods). We achieved this goal through probabilistic topic modeling^14^, originally proposed for learning word co-occurrence patterns in documents with different semantic topics. With this approach (see details in Methods), we were able to identify ten distinct “PASC topics”, each of which is characterized by a unique post-acute infection incidence probability distribution across the 137 individual conditions.

Figure 2 shows the heatmap matrix of the learned topics from the INSIGHT cohort. Each column was a learned topic, and each row was a potential PASC condition category (we demonstrated 31 of them in the heatmap and aggregated the remaining 106 because none of their incident probabilities exceeded 0.1 in any of the learned topics). Each entry in the matrix corresponded to the probability of the specific PASC condition in the corresponding topic. All entry values in the same column added up to one so that each topic was characterized by a rigorous incident probability distribution over the 137 PASC conditions. Specifically, Topics T1, T2, and T5 concentrated on the conditions of the musculoskeletal system, digestive system, and nervous system, respectively. T4, T7, and T9 included respiratory conditions mixed with sleep disorder and anxiety along with symptoms such as headache and chest pain. T3 included fluid and electrolyte disorders combined with anemia and cardiac problems. T6 was musculoskeletal and skin conditions with headache and fatigue problems. T8 was anemia and digestive system problems. T10 was a mixture of circulatory problems, renal failure, fluid and electrolyte problems, and others.

**Figure 2.**
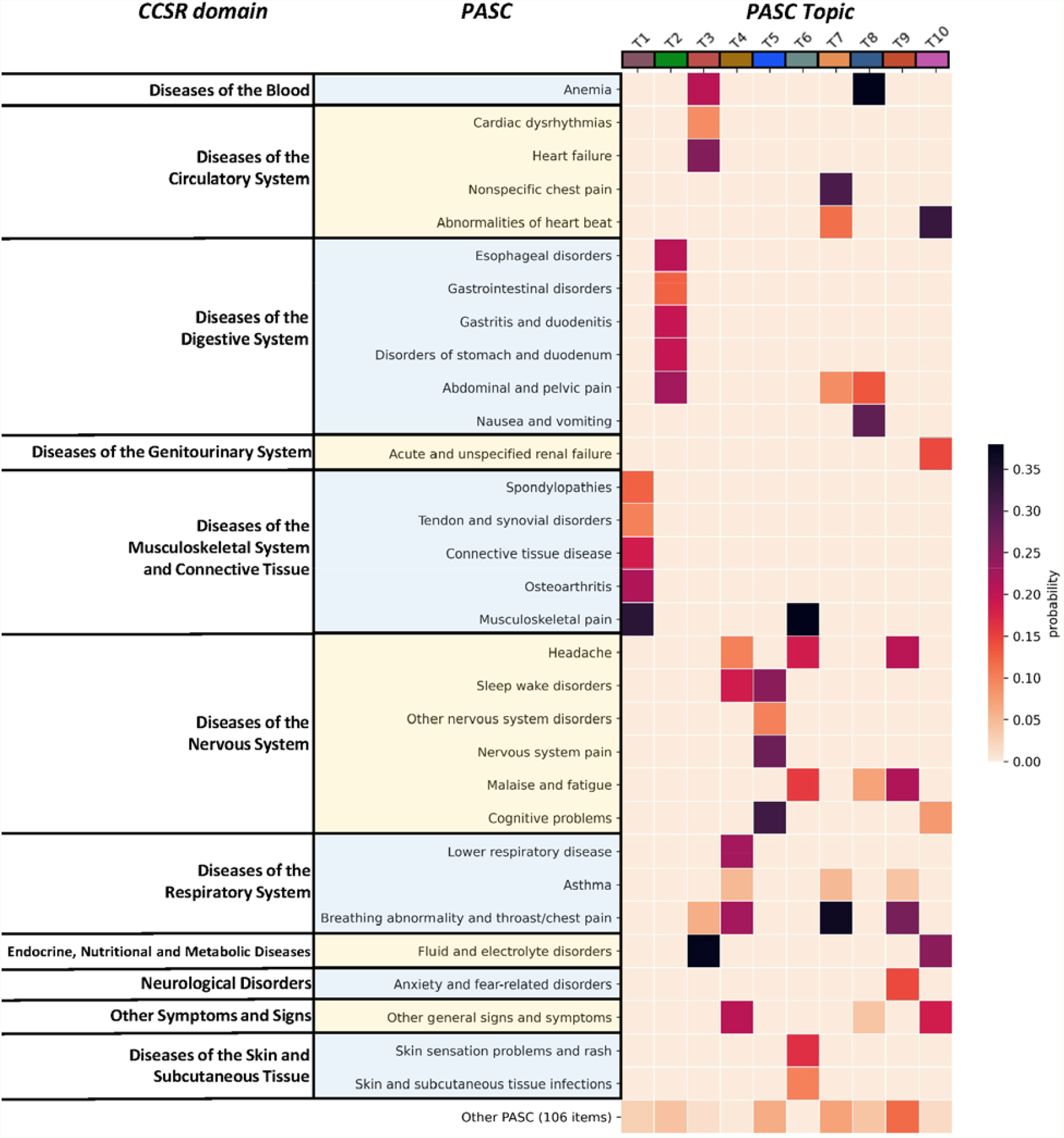
The heatmap of PASC topics learned on the INSIGHT cohort. Each row denotes a potential PASC category grouped by different CCSR domains, and each column denotes a particular PASC topic. Each PASC topic is characterized by a unique post-acute incidence probability distribution over all 137 individual potential PASC categories.

### Potential PASC Subphenotypes

With the potential PASC topics identified, we could describe PASC-affected patients with them and derive potential PASC subphenotypes as patient clusters (Methods). In the INSIGHT cohort, four subphenotypes were identified. Table 2 summarizes their characteristics with respect to patient demographics, disease severity in the acute phase according to treatment setting, as well as the prevalence of comorbidities in the baseline period. Across the subphenotypes, we also demonstrated the prevalence of potential PASC conditions and incident prescriptions of medications in the post-acute infection period for patients in different subphenotypes in Figures 3 and 4. From Figure 3, we observed that four subphenotypes had different prevalent PASC conditions, which were consistent with the top PASC conditions in the topics with large proportions. Next, we characterized these subphenotypes in detail as follows.

**Table 2.**
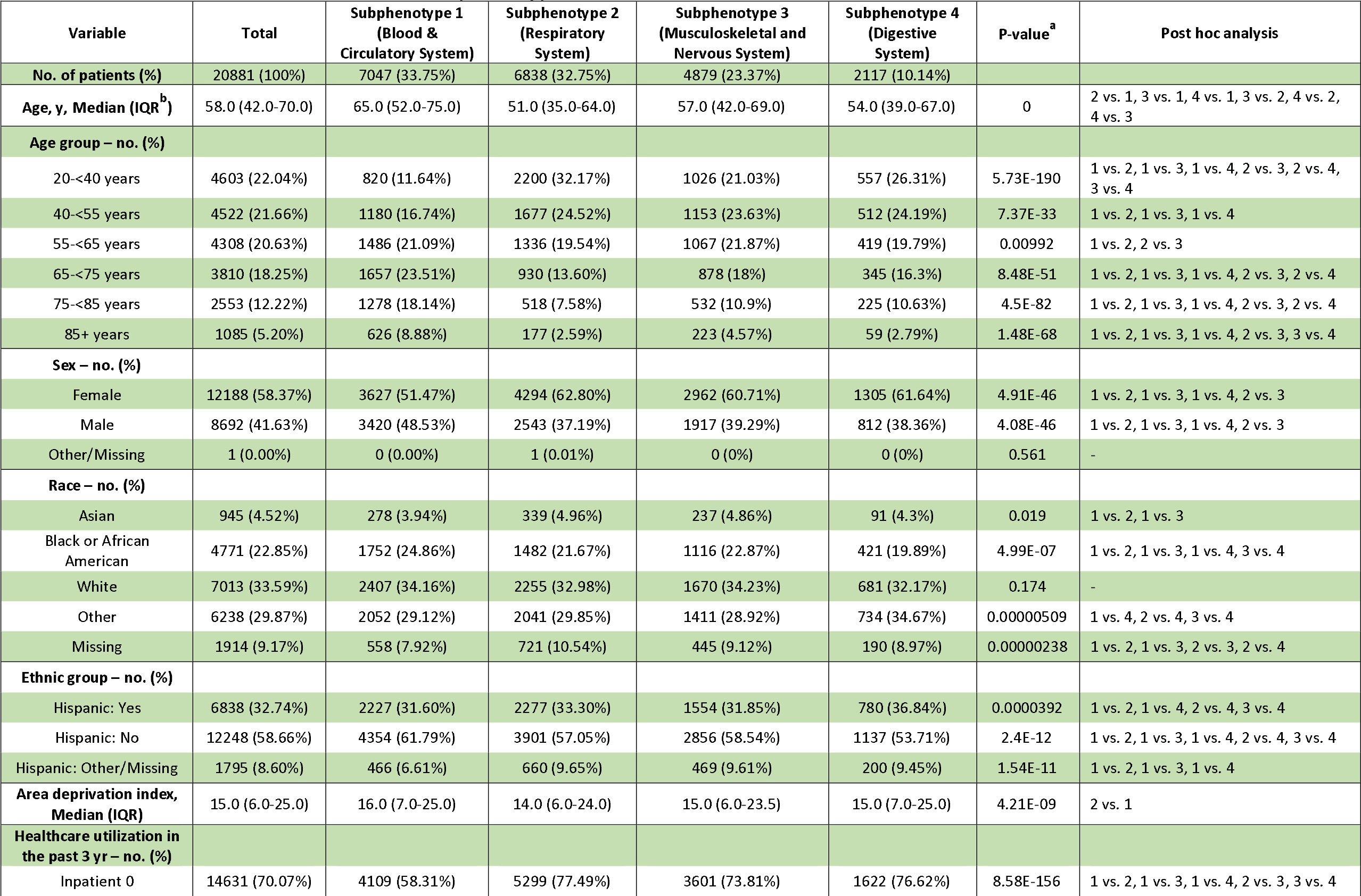
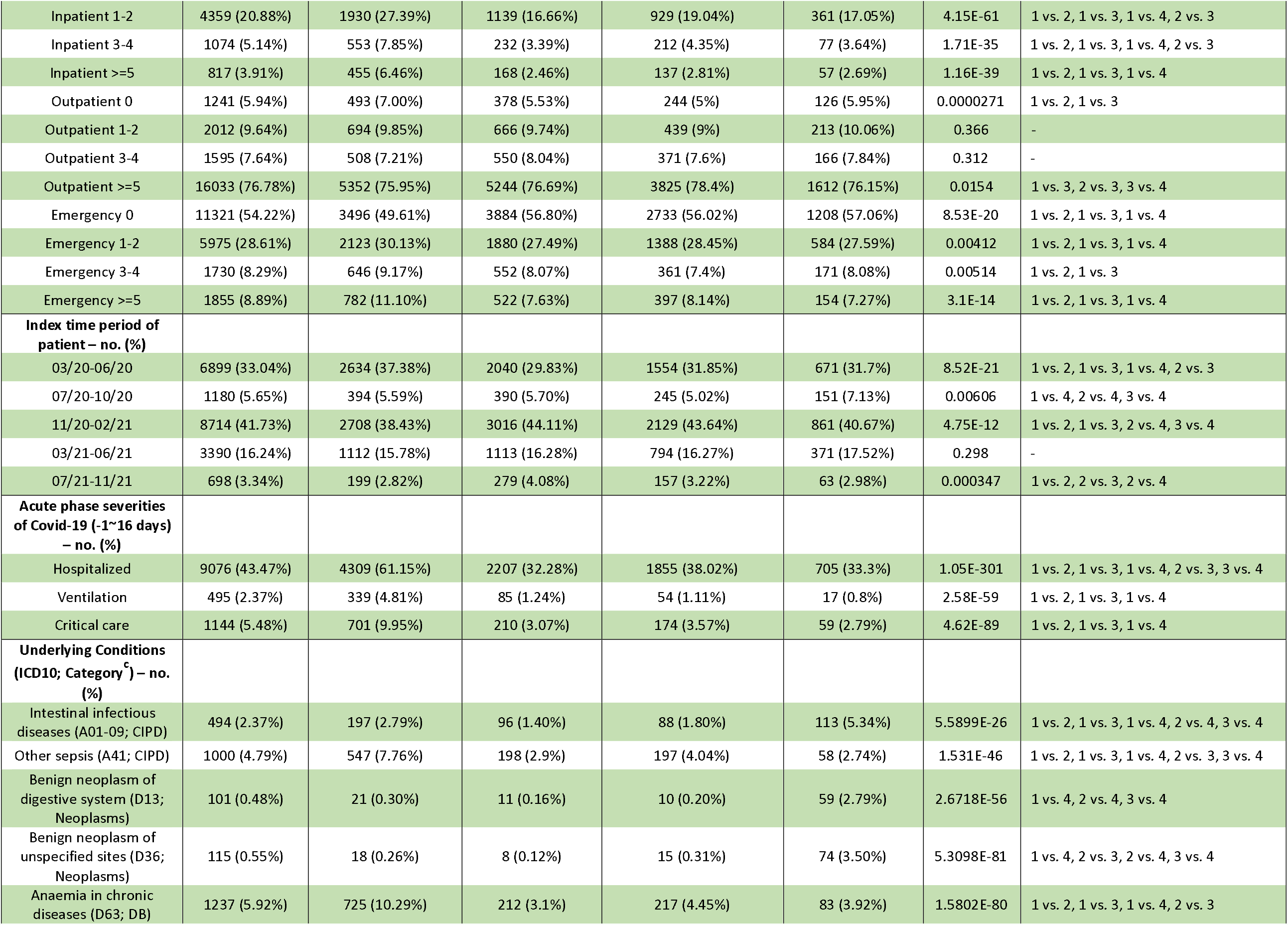
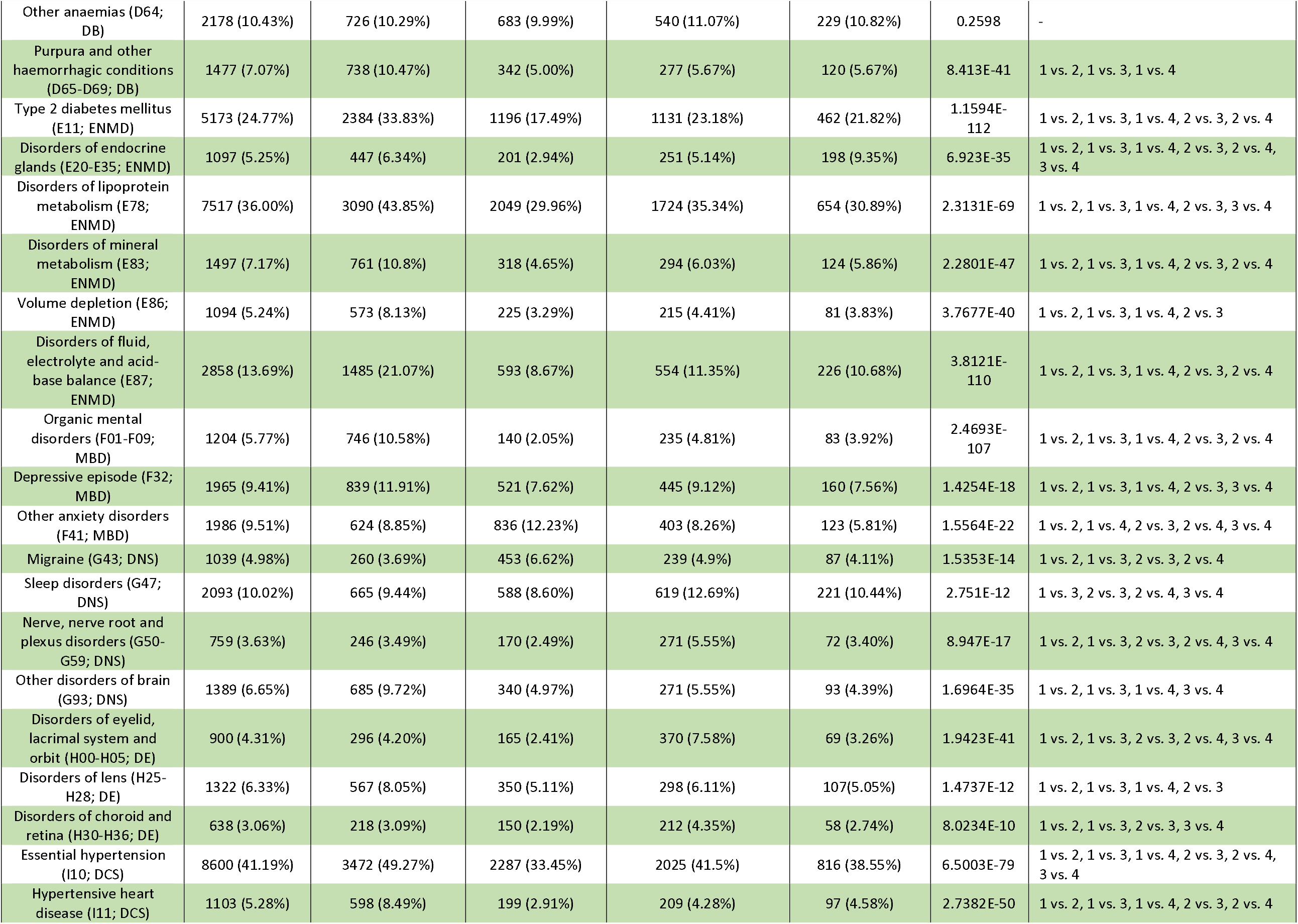
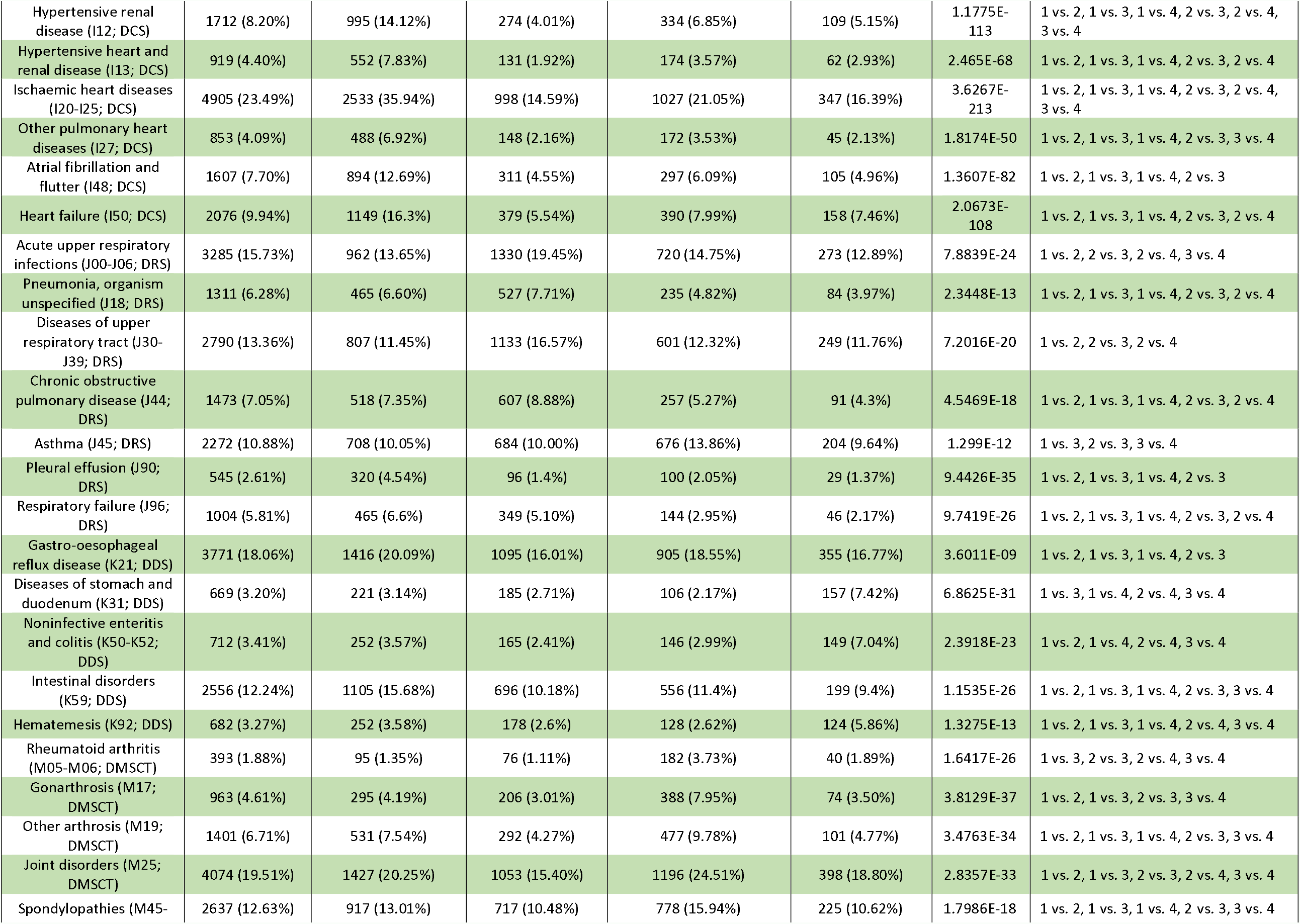
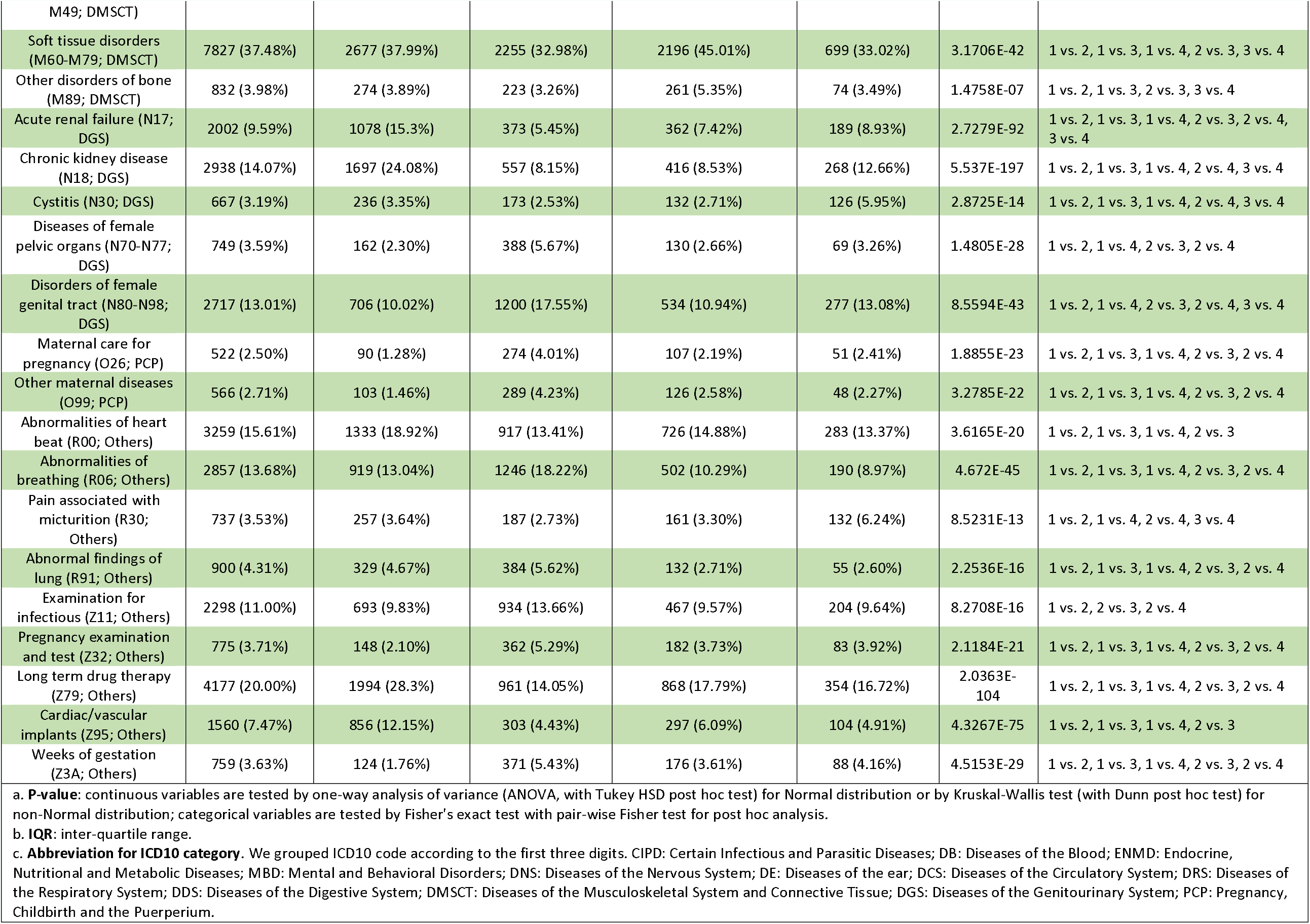
Characteristics of the identified subphenotypes on the INSIGHT cohort.

**Figure 3.**
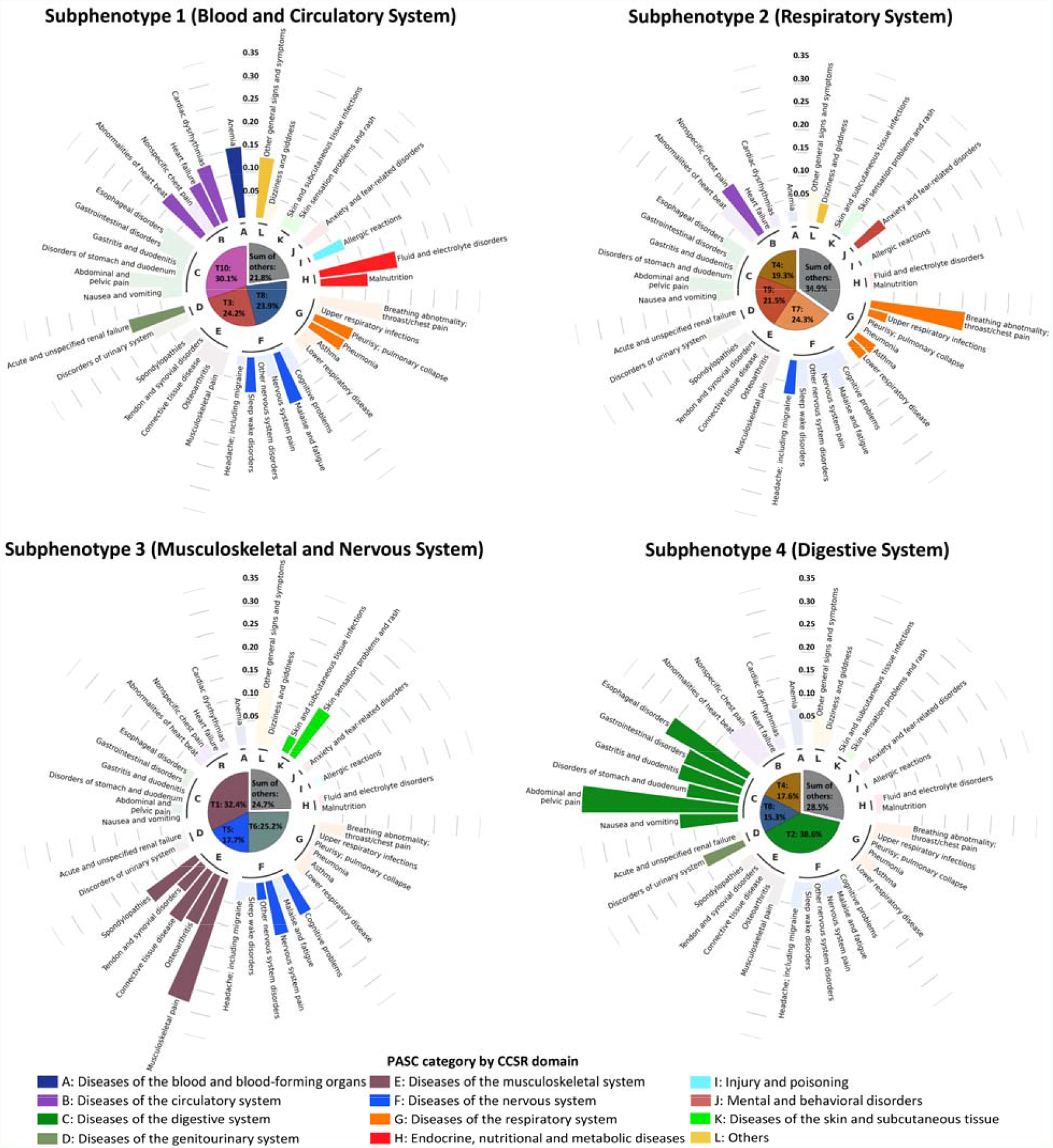
The incidence rates of potential PASC conditions in each subphenotype for the INSIGHT cohort, where potential PASC conditions were grouped into different categories shown in different colored bars outside the center pie chart. A condition is highlighted in the subphenotype where it has the highest incidence rate. The center pie chart of each subphenotype shows the mean topic proportions of the patients it included, and the meanings of the topic indices can be referred to Figure 2.

**Figure 4.**
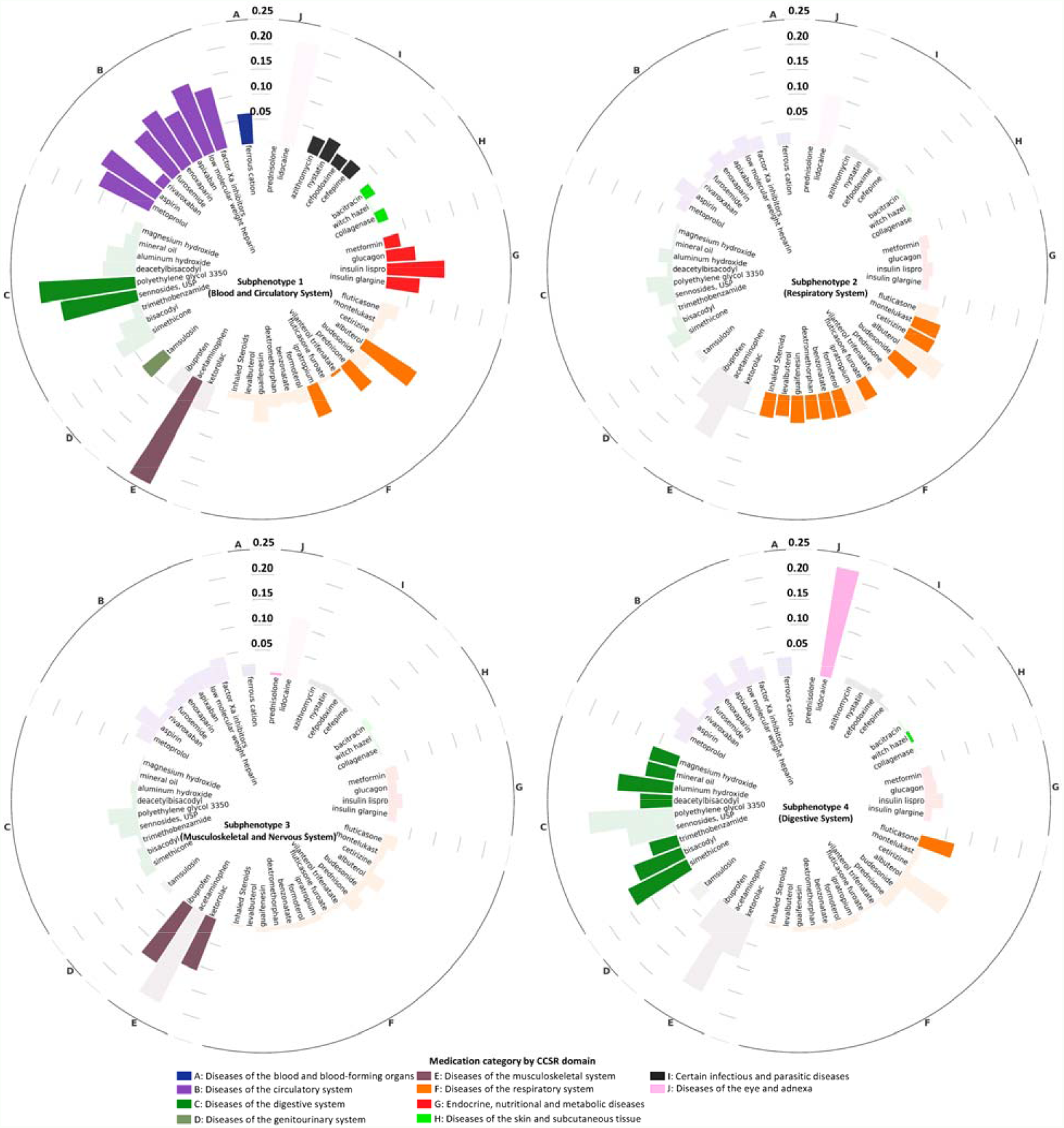
The prevalence of incident prescriptions of medications in the post-acute infection period for each subphenotype on the INSIGHT cohort, where medications are grouped into different categories shown by different colors. For one of the medications, if it is most prevalent in one subphenotype, we highlighted it in this subphenotype.

#### Subphenotype 1 (Blood and Circulatory System)

consisted of 7,047 (33.75%) patients. It was dominated by blood- and circulation-related topics (T3, T8, T10), including anemia, fluid and electrolyte problems, and circulatory and cardiac problems. Compared to other subphenotypes, patients in this subphenotype were older (median age 65.0 years, IQR [52.0- 75.0]) and had the highest proportion of males (48.53%). They also had a higher severity of COVID-19 in the acute phase (with the highest rate of hospitalization [61.15%], use of mechanical ventilation [4.81%], and critical care services [9.95%]). Furthermore, this subphenotype had the highest portion of patients (37.38%) infected with SARS-CoV-2 during the first wave of the pandemic (March to June 2020) in NYC. In addition, patients in this subphenotype had a higher prevalence of underlying conditions than other subphenotypes, especially for blood, circulation, and endocrine comorbidities. Correspondingly, patients in this subphenotype had a high incident prescription for medications to treat circulatory and endocrine problems and anemia.

#### Subphenotype 2 (Respiratory System)

included 6,838 (32.75%) patients. It was dominated by respiratory conditions (topics T4, T7, and T9), sleep disorders, anxiety, and symptoms such as headache and chest pain. This subphenotype had the youngest patients among the four (median age 51.0 years, IQR [35.0-64.0]), the highest proportion of females (62.8%), and the lowest rate of hospitalization (31.28%) for COVID-19. It also had the largest proportion of patients who tested positive for SARS-CoV-2 from November 2020 to November 2021 (64.47%). Patients in this subphenotype had higher baseline comorbidity burdens for respiratory conditions such as upper respiratory problems and chronic obstructive pulmonary disease, breathing problems, and a higher incident prescription for a diverse set of anti-asthma, anti-allergy, and anti-inflammation medications including inhaled steroids, levalbuterol, and montelukast.

#### Subphenotype 3 (Musculoskeletal and Nervous System)

consisted of 4,879 (23.37%) patients. It mainly contained musculoskeletal and nervous system problems (topics T1, T5, and T6) such as musculoskeletal pain, headaches, and sleep-wake problems. This subphenotype included patients with a median age of 57.0 years (IQR [42.0-69.0]), with 60.71% female. It had the highest proportion of patients with more than five outpatient visits (78.4%). Patients in this subphenotype had higher baseline comorbidity burdens of autoimmune and allergy conditions such as rheumatoid arthritis and asthma, as well as other musculoskeletal and nervous system problems including soft tissue, bone, and sleep problems. This subphenotype was also associated with a higher incident prescription risk of pain medications (ibuprofen and ketorolac) in the post-acute infection period.

#### Subphenotype 4 (Digestive System)

included 2,117 (10.14%) patients mainly with digestive system problems such as abdominal pain, vomiting, and respiratory conditions (topics T2, T4, T8). Patients in this subphenotype had a median age of 54.0 (IQR [39.0-67.0]) with 61.64% female. Patients in this subphenotype had the highest proportion of patients without any baseline emergency visits (57.06%) and the lowest rates of mechanical ventilation (0.8%) and critical care admission (2.79%) in the acute phase of COVID-19. Compared with the other three subphenotypes, this subphenotype had an overall lower prevalence of underlying conditions, and a slightly higher prevalence of digestive problems such as hematemesis, stomach and duodenum disorders, and digestive system neoplasm. In addition, this subphenotype had higher incident prescription rates of drugs for treating the digestive system.

### Contrast with COVID-19 Negative Patients

These potential PASC subphenotypes were derived from SARS-CoV-2 infected patient. However, it was unclear how the potential PASC diagnosis co-incidence patterns encoded in them differed from the non-infected patients. To answer this question, we compared the incidence patterns of 28 selected potential PASC conditions in 30-180 days after the COVID-19 lab test between positive and matched negative patients (Methods). The results were demonstrated in Figure 5, where the nodes in each network corresponded to a particular potential PASC condition with their sizes proportional to the incidence rate in the corresponding subphenotype or matched controls, and each line linking a pair of nodes indicated co-incidence of the corresponding potential PASC diagnoses with its thickness proportional to the co-incidence rate. From the figure, we could see that the conditions we used to characterize each subphenotype were clearly associated with larger-sized nodes, representing higher incidence rates. At the same time, we do not observe differences in node sizes on the matched COVID-19 negative networks. In addition, we observed denser connections in COVID-19 positive networks, which suggested that the potential PASC conditions did not appear independently, but collectively, and those larger nodes were network hubs associated with more lines.

**Figure 5.**
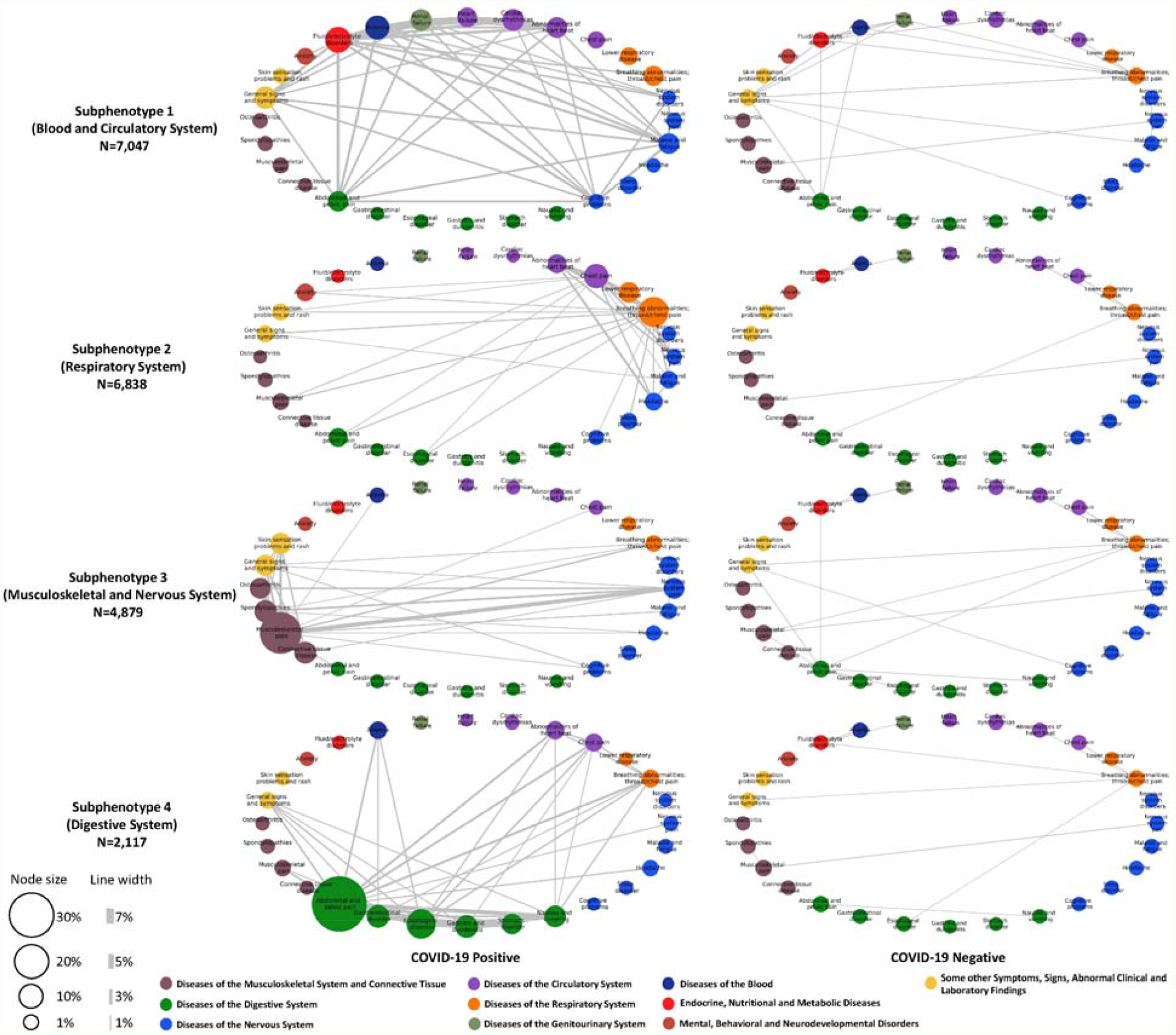
Difference of the incidence patterns of selected PASC conditions (grouped by CCSR domains) in 30-180 days after COVID-19 lab test between positive and matched negative patients on the INSIGHT cohort. The bubbles in each network correspond to a PASC condition with their sizes proportional to the incidences in the particular subphenotype or matched controls. The edge linking a pair of bubbles indicates co-incidence of the corresponding potential PASC conditions with its thickness proportional to the co-incidence rate, where lines are visible if the rate is larger than 1%.

### Replication on OneFlorida+ CRN

We repeated the same subphenotyping process on the OneFlorida+ cohort; the subphenotypes generated were highly overlapping with those from the INSIGHT cohort. Following the same analytical pipeline (Figure 1), we first identified incident diagnoses for the 137 potential PASC conditions with 30-180 days of testing positive for SARS-CoV-2; we then conducted topic modeling. Supplemental Figure 6 displays the heatmap of all learned potential PASC topics, where we see topics concentrated on problems with the musculoskeletal system (T1), digestive system (T2), nervous system (T5), and topics mixed with respiratory system problems and blood/circulatory system problems (T3), as well as headache and sleep-wake problems (T7). Some topics also include a mixture of diagnoses. For example, T9 is throat/chest pain mixed with breathing/heartbeat abnormalities; T6 is a mixture of musculoskeletal pain, headache, malaise and fatigue, and skin sensory problems; T8 and T10 are topics mixed with electrolyte/fluid disorders and anemia/arrhythmias, and T4 is a topic mixing up problems involving digestive, nervous and respiratory systems. To quantify areas of overlap with findings from the INSIGHT cohort, we evaluated the pairwise similarities between the topics learned from the two different cohorts (Methods) and visualized the results in Supplemental Figure 4, which clearly showed a one-to-one correspondence between the topics learned from the two cohorts (as identified by darker colors on the diagonal line).

With the learned topics, we also built topic loading-based representations for patients in the OneFlorida+ cohort, derived four potential PASC subphenotypes with HAC (Methods), and described the characteristics of patients classified into each subphenotype. (Supplemental Table 3, Figures 7 and 8). Subphenotype 1 was dominated by incidental blood and circulatory system problems in the post-acute infection period, which included 25.43% of the patients who were older (with a median age of 62.0 and IQR [49.0-74.0]), with the highest proportion of males (46.93%, compared to 38.29% for the overall population) and the highest rates of hospitalization (57.34%, compared to 36.69% for the overall population), mechanical ventilation (8.57%, compared to 3.39% for the overall population) and critical care admission (12.52%, compared to 6.07% for the overall population) in the acute phase of COVID-19. This subphenotype was associated with a higher prevalence of underlying conditions and more new prescriptions for medications treating circulatory system, blood, and endocrine problems. Subphenotype 2 was dominated by incidental respiratory problems and was the largest subphenotype containing 5281 (38.48%) patients with a median age of 47.0 (IQR [33.0-61.0]). They had a higher prevalence of respiratory conditions at baseline including chronic obstructive pulmonary disease, pneumonia, upper respiratory tract problems, and had a higher post-acute infection incident prescription for respiratory medications. Subphenotype 3 was dominated by incident problems with musculoskeletal and nervous problems in the post-acute infection phase. It included 3205 (23.35%) patients with a median age of 48.0 (IQR [33.0-61.0]) and had the lowest hospitalization rate in the acute phase (27.8%). This subphenotype had a higher prevalence of baseline musculoskeletal and connective tissue problems and asthma, and more new prescriptions for pain medications, including ketorolac and ibuprofen in the post-acute infection phase. Subphenotype 4 was dominated by incidental digestive problems. It was the youngest (median age 46.0 [32.0-60.0]) and smallest (including 1748 [12.74%] patients) subphenotype, with the highest proportion of females (67.11%, compared to 61.70% overall) and lowest rates of mechanical ventilation (0.97%) and critical care admission (2.8%) in the acute phase. This subphenotype was associated with a higher baseline burden of digestive system problems, and more new prescriptions for medications focused on the digestive system. These observations and characterizations were highly consistent with the subphenotypes identified from the INSIGHT cohort. In addition, the difference in the incidence patterns of 28 selected potential PASC conditions in 30-180 days after the COVID-19 lab test between positive and matched negative patients on the OneFlorida+ cohort is shown in Supplemental Figure 9, which was also highly consistent with the results from the INSIGHT cohort.

## Discussion

Many studies have pointed out the existence of a diverse set of symptoms and signs that may develop, persist, or recur in the post-acute SARS-CoV-2 infection period^7^. These conditions, typically referred to as PASC, involve a wide range of organ systems. Different from most of the existing research which studied these conditions independently, we developed a data-driven framework shown in Figure 1 to identify subphenotypes of SARS-CoV-2 infected patients based on newly incident symptoms and signs from 30 to 180 days (the post-acute period) after their infection confirmation, such that patients within the same subphenotype share a similar distribution of potential PASC condition incidences in the post-acute periods.

Within both INSIGHT and OneFlorida+ CRNs, we identified four consistent subphenotypes dominated by new conditions of the blood and circulatory systems (Subphenotype 1), respiratory system (Subphenotype 2), musculoskeletal and nervous systems (Subphenotype 3), and digestive system (Subphenotype 4).

Comparing the subphenotypes derived from both cohorts, we observed that Subphenotype 1 included older patients with higher baseline comorbidity burden, with greater severity of illness during the acute phase (e.g., higher rates of hospitalization, critical care admission and mechanical ventilation) and with higher incident prescription rates of medications for treating diseases of many different organ systems. This subphenotype had the highest proportion of males, which aligns with the finding that males had more severe acute SARS-CoV-2 infections^15^. This subphenotype had a large proportion of patients with their SARS-CoV-2 infection confirmed during the early pandemic (March to September 2020) when treatment standards were still evolving. Temporally, NYC was the epicenter for the first wave. This may explain the observation that this was the largest subphenotype for INSIGHT (containing 33.75% of the patients) but the second largest subphenotype for OneFlorida+ (containing 25.43% of the patients). Early cases had greater acute phase severity, which may explain the more severe incident conditions (e.g., heart failure and renal failure) in the post-acute infection period of these patients, which could be caused by the hyperinflammation during the acute phase^16^.

Subphenotype 2 is another major subphenotype for both cohorts (the second largest for INSIGHT occupying 32.75% of the patients, and the largest for OneFlorida+ accounting for 38.48% of the population). It is the youngest subphenotype for INSIGHT and the second youngest subphenotype for OneFlorida+ and contains the highest proportion of patients who had an acute SARS-CoV-2 infection confirmed from July to November 2021. The young age and infection recency are consistent with the milder incident PASC conditions of this subphenotype. Of note, patients in this subphenotype had a high baseline rate of pulmonary comorbidities which is likely correlated with the high rate of incident respiratory medication prescriptions in the post-acute SARS-CoV-2 period.

Subphenotype 3 and 4 were two smaller subphenotypes with Subphenotype 3 associated with musculoskeletal and neurological conditions, whereas Subphenotype 4 was associated with gastrointestinal conditions. Patients in Subphenotype 3 also suffered from dermatologic conditions and had the highest rates of related conditions at baseline, including autoimmune diagnoses such as rheumatoid arthritis and allergy conditions. Conversely, patients in Subphenotype 4 had the mildest acute phase severity (e.g., lowest rates of mechanical ventilation and critical care admissions.

There are several strengths in our study. First, we adopted a topic modeling approach to derive compact patient representations based on the co-incidence patterns across different diagnoses. Unlike other dimensionality reduction techniques such as Principal Component Analysis (PCA)^17^, topic modeling is designed specifically for data samples with binary or count features^18,19^ and thus appropriate for our analysis. Second, INSIGHT and OneFlorida+ include patients from distinct geographic regions in the US with different characteristics, allowing us to validate the robustness of the derived subphenotypes. Third, our study period (March 2020 to November 2021) covers different COVID-19 waves associated with different SARS-CoV-2 virus variants. Our study cohorts contained robust patient populations in New York and Florida, representing the different waves of SARS-CoV-2 infected cases in US. This is important factor contributing to the different distributions of the four subphenotypes in the two cohorts.

Our study is not without limitations. First, our analysis is based on longitudinal observational patient data, which cannot explain the biological mechanisms behind PASC directly. Second, the PASC diagnoses we investigated were encoded as CCSR categories, which may not reflect the co-incidence patterns of fine-grained diagnosis conditions in the context of PASC. Third, we focused on new incidences of conditions in the post-acute infection period for COVID-19 patients and did not consider pre-existing conditions that are persistent or exacerbated due to the acute SARS-CoV-2 infection. Finally, our study period did not represent the recent wave dominated by the Omicron variants of SARS-CoV-2.

To summarize, our study dissects the complexity and heterogeneity of newly incident conditions in 30-180 days after SARS-CoV-2 infection confirmation into four reproducible subphenotypes based on the EHR repositories from two large CRNs using machine learning. These four subphenotypes included a severe one involving problems with the blood and circulatory system and associated with high baseline comorbidity burden and disease severity in its acute phase, a milder one in younger people mainly with respiratory problems, and two pain-dominated ones (musculoskeletal/nervous system pain and abdominal pain respectively). Overall, patients in each subphenotype tend to have higher rates of related conditions in the baseline period. Our study provides the first systematic study on the co-incidence patterns of conditions in the post-acute infection period of SARS-CoV-2 infected adult patients, which can inform more nuanced and tailored diagnosis and treatment plans.

## Methods

### EHR Data Repositories

Two large-scale de-identified real-world EHR data warehouses were utilized in our analyses. Our first cohort data was based on the EHR from INSIGHT CRN^9^, which contains the longitudinal clinical information of around 12 million patients in New York City area. Our second cohort data was based on the EHR from the OneFlorida+ CRN^10^, which contains the information of nearly 15 million patients majorly from Florida and selected cities in Georgia and Alabama. The use of the INSIGHT data was approved by the Institutional Review Board (IRB) of Weill Cornell Medicine following protocol 21-10-95-380 with title “Adult PCORnet-PASC Response to the Proposed Revised Milestones for the PASC EHR/ORWD Teams (RECOVER)”. The use of the OneFlorida+ data for this study was approved under the University of Florida IRB number IRB202001831.

### Potential PASC Conditions

We compiled a list of 137 diagnostic categories covering near 6,500 ICD-10-CM codes as potential PASC conditions for our study. The list was built based on the Clinical Classifications Software Refined (CCSR) v2022.1 covering 66,534 ICD-10-CM Diagnoses. The codes that would not plausibly be considered post-acute sequelae of COVID-19 in the adult population (e.g., HIV, tuberculosis, infection by non-COVID causes, neoplasms, injury due to external causes, etc.) were excluded, and parent codes (e.g., the first 3-digits of ICD-10 codes) were systematically added. The full list of diagnosis codes for the PASC is provided in Supplemental Table 1.

### Cohort Construction

For the both cohorts, adult patients (age ≥ 20) with at least one SARS-CoV-2 polymerase-chain-reaction (PCR) or antigen laboratory test (Supplemental Table 2) between March 01, 2020 and November 30, 2021 were selected. Then we chose the patients who had at least one positive test and had at least one potential PASC conditions in the follow-up (or post-acute infection) period defined as below. We further made sure those potential PASC conditions were new incidences in the follow-up period by excluding patients who had any of them in both baseline and follow-up periods. The overall inclusion-exclusion cascade was shown in Supplemental Figure 1, and the relevant definitions are provided below.

- Index date: the date of the first COVID-19 positive test.
- Baseline period: from 3 years to one week prior to the index date.
- Follow-up (post-acute infection) period: from 31 days after the index date to the day of documented death, last record in the database, 180 days after baseline, or the end of our observational window (Nov. 30, 2021), whichever came first.

### Topic Modeling

We use binary vectors 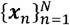 to represent the patients, where *n* is the patient index the *i*-th element of ***x***_*n*_, or ***x***_*n*_(*i*) =1 if the *i*-th potential PASC condition appears in the post-acute infection period of the *n*-th patient’s EHR, otherwise ***x***_*n*_(*i*) = 0. Therefore, each is a 137-dimensional binary vector (Step 1 in Figure 1). Topic modeling (TM)^14^ as then applied on these vectors to learn a set of potential PASC topics. Specifically, assume that each patient can be represented as a mixture of *K* latent PASC topics Φ ∈ ^*RD* × *K*^, where each topic ***φ***_*k*_ can be viewed as a set of PASC that are more likely to be co-incident in the post-acute infection period of a particular patient (Step 2 in Figure 1). Then for each patient, TM infers the mixture memberships ***θ****n* ∈ *R*^*K*^, also called topic proportions or topic loadings, as the new representation for each patient (Step 3 in Figure 1). A patient with higher loading value on a particular topic indicates that he/she suffers from more co-incident condition patterns from this topic. In other binary space ***x***_*n*_ to the low-dimensional continuous PASC topic space ***θ****n*, which will be words, TM transforms the representations of each patient from the original 137-dimensional leveraged further for subphenotype identification through clustering later (Step 4 in Figure 1). Specifically, we used Poisson factor analysis (PFA)^20^ as the concrete TM method, which generates ***x***_*n*_ as follows.

- Draw a topic proportion ***θ***_*n*_ for the *n*-th patient from a Gamma distribution ***θ***_*n*_ *Gamma* (1,1);
- Draw the *k*-th PASC topic from a Dirichlet distribution ***φ***_*k*_ ∼ *Dirichlet* (0.01), *k* = 1, …, *K*;
- Draw a binary vector ***x***_*n*_ from the Bernoulli distribution by Bernoulli-Poisson link^21^:

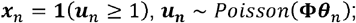

where 1(·) is an indicator function representing ***x***_*n*_ =1 if ***u***_*n*_ ≥ 1, and ***x***_*n*_ =0 if ***u***_*n*_ = 0. We use Gibbs sampling^22^ to infer the posterior of PASC topic **Φ** and topic proportions 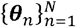.

### Determining the Number of Topics

The number of topics, *K*, is an important parameter in TM. To determine an optimal *K* based on the data, we used two metrics: data likelihood and topic coherence^23^. Data likelihood is used to evaluate the fitness of the model on the current data set, where the larger value indicates better fitness. Topic coherence is used to evaluate the relevance of our learned PASC topics to the investigative condition list, where the value is from zero to one and higher value indicates better coherence. Detailed calculations of these two metrics are provided in the Supplemental methods and results are shown in Supplemental Figure 2. From this figure, we can see that more topics can provide higher data likelihood because we have more topics to represent the original PASCs. However, more topics may bring down topic coherence, which suggests the redundancy between the newly added and the old ones. With these considerations, we set the final number of topics as 10 for both INSIGHT and OneFlorida+ cohorts as it achieved the best topic coherence and reasonable data likelihood (we do not want the data likelihood to be too perfect as that may suggests overfitting).

### Topic Robustness

As TM is a probabilistic process, we want to guarantee the robustness of the identified topics. To achieve this goal, we firstly did 1000 bootstrapping (randomly choose 80% patients from each cohort) to learn topics. Then according to the importance of each topic (mean topic loadings over all patients), we reordered the topics and calculated the cosine similarity among all topics from each pair of bootstrapping:

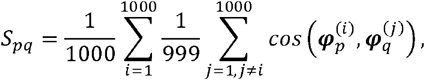

where 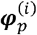 is the *p*-th topic vector learned from the -th bootstrapped samples, and *s*_*pq*_ is the similarity between the *p*-th topic and the *q*-th topic. Supplemental Figure 3 demonstrated the heatmap of the similarity matrix with *spq* as its (*p, q*) -th entry, from which we can clearly observe a darker diagonal line, which indicates a high similarity between the topics learned from different bootstrapped samples and thus implies the learned topics are robust.

### Topic Consistency

We also quantitatively evaluated the consistency between the two set of topics learned from different cohorts. Specifically, denoting the topic matrices learned from the two cohorts as **Φ**^**;(1)**^ and **Φ**^**;(2)**^, then we can evaluate the consistency between the *i*-th topic in cohort 1 and the *j*-th topic in cohort 2 as the cosine similarity between their corresponding topic vector 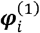 and 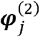 as:

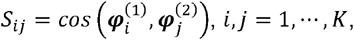

Finally, the heatmap of the topic consistency matrix with *s*_*ij*_ as its (*i,j*) -th entry was shown in Supplemental Figure 4, from which we can clearly observe darker diagonal values, which suggests high consistency between the two sets of topics.

### Subphenotyping through Clustering

With the learned *K*-dimensional topic loading vector for *n*-th patient ***θ***_*n*_, we applied hierarchical agglomerative clustering method with Euclidean distance calculation and Ward linkage criterion^24^ to derive subphenotypes as patient clusters. For determining the optimal number of clusters (subphenotypes), we applied NbClust R package^25^, which includes 21 cluster indices to evaluate the quality of clusters. With the patients from the INSIGHT and OneFlorida+ CRN, 13 and 12 out of the 21 indices agreed 4 is the optimal number of clusters. Through majority voting, we set the number of clusters as 4 in both two cohorts. Supplemental Figure 5 demonstrates the UMAP embeddings^26^ and dendrogram of these clusters for both cohorts.

We have also examined the robustness of the identified clusters on both cohorts. Specifically, for each cohort, we used the subphenotypes derived from all patients as references, so that the subphenotype index for the *n*-th patient is denoted as *y*_*n*_. Then we ran 1000 bootstrapping (randomly choose 80% patients from the cohort denoted as set Ωi,*i* = 1,, 1000) to learn topic model and then derive subphenotypes with the same procedure as described above. For each bootstrapped sample set *i*, we can obtain another subphenotype index for *n*-th patient from Ωi as *ŷ*_*n,i*_. We calculated mean and 95% confidence interval of adjusted rand score (ARI) and normalized mutual information (NMI) between the clustering results on bootstrapped sample sets and reference as 0.902 (95% confidence interval (CI): 0.863 − 0.927) and 0.937 (95% CI 0.908 − 0.952) for the INSIGHT cohort, and 0.914 (95% CI: 0.907 − 0.929) and 0.950 (95% CI: 0.936 − 0.968) for the OneFlorida+ cohort, which suggests the identified clusters are highly robust.

### Comparison with SARS-CoV-2 Infection Negative Patients

We compared the co-incidence patterns of the investigative conditions in the follow-up periods for patients with SARS-CoV-2 infection testing positive and negative. The SARS-CoV-2 infection negative patients are with all negative results for their SARS-CoV-2 infection lab tests during March 2020 to November 2021, and there were no documented COVID-19 related diagnoses during this time period. The index date for each individual patient in non-infected group is defined as the date of the first (negative) lab test.

To make fair comparisons, we performed similarity matching to identify appropriate negative patients for each positive patient based on the following hypothetical confounding variables.

- Demographics: age, gender, race, and ethnicity, where age was binned into different groups (20-<40 years, 40-<55 years, 55-<65 years, 65-<75 years, 75-<85 years, 85+ years).
- The area deprivation index (10-rank bins of national ADI) for capturing socioeconomic disadvantage of patients’ neighborhood^13^.
- Index date for considering the effect of different stages of pandemic, which was binned into different time intervals (March 2020 – June 2020, July 2020 – October 2020, November 2020 - February 2021, March 2021 – June 2021, July 2021 – November 2021).
- Medical utilizations measured by numbers of inpatient, outpatient, and emergency encounters in the baseline period (binned into 0 visit, 1 or 2 visits, 3 or 4 visits, 5+ visits for each encounter type).
- Coexisting conditions including comorbidities and medications based on a tailored list of the Elixhauser comorbidities^27^. We defined the patient having a particular condition if he/she had at least two related records during the baseline period.

For identifying the negative controls for each patient in a particular subphenotype, we first required exact match for confounders of demographics, ADI, and index date to obtain an initial set, and then performed robust propensity score (PS) matching on other hypothetical confounders robust propensity score to rank the patients in the initial set and we finally picked the top 2. We used standardized mean difference (SMD) to quantify the goodness-of-balance of confounders between two groups 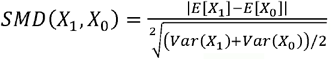, where *SMD* < 0.2 is the threshold to examine whether this confounder is balanced^28^. On both ISNIGHT and OneFlorida cohort, we found that all confounders on all subphenotypes were balanced.

## Data Availability

All data produced in the present study are available upon reasonable request to the corresponding authors

## Code availability

For reproducibility, our codes are available at https://github.com/haozhangWCM/Subphenotyping-for-PASC. We used Python 3.7, python package scikit-learn-0.23.2, numpy-1.16.5, umap-learn-0.5.1, and scipy-1.7.3 for machine learning models.

## Data availability

The INSIGHT data can be requested through https://insightcrn.org/. The OneFlorida+ data can be requested through https://onefloridaconsortium.org. Both the INSIGHT and the OneFlorida+ data are HIPAA-limited. Therefore, data use agreements must be established with the INSIGHT and OneFlorida+ networks.

## Acknowledgement

This research was funded by the National Institutes of Health (NIH) Agreement OTA OT2HL161847 (contract number EHR-01-21) as part of the Researching COVID to Enhance Recovery (RECOVER) research program.

## Author contributions

H.Z and F.W. proposed the initial idea. H.Z. designed and implemented the framework and analyzed the results. C.Z and J.X. preprocessed the INSIGHT and OneFlorida dataset, respectively, and helped to analyze the results. Z. X. did statistical analysis. All the authors contributed to the final writing of the paper.

## Supplemental Figure and Table

**Supplemental Table 1**. PASC list in our study defined by CCSR domain with corresponding ICD-10-CM codes.

**Supplemental Table 2**. SARS-CoV-2 polymerase-chain-reaction or antigen laboratory test.

**Supplemental Table 3.**
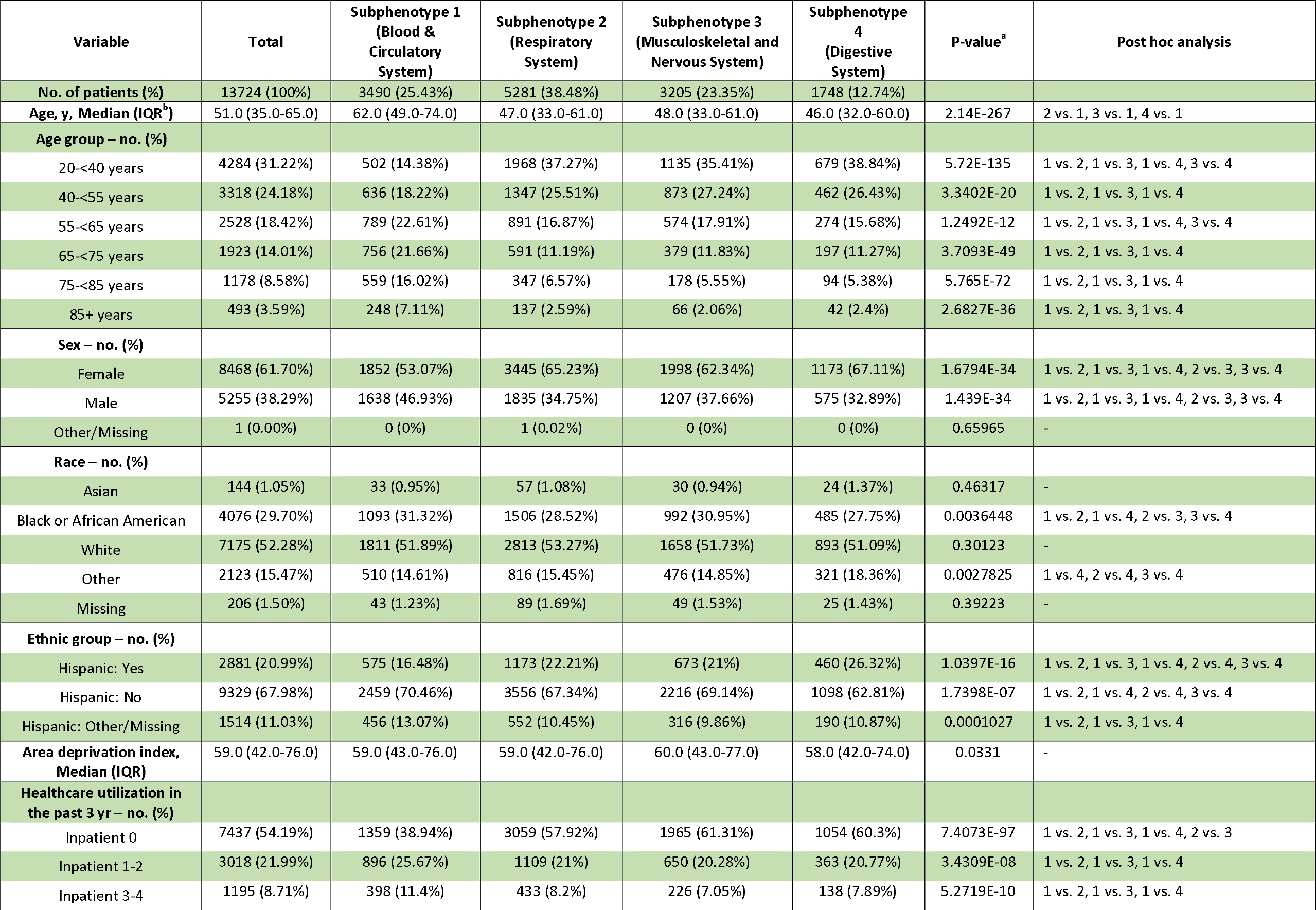
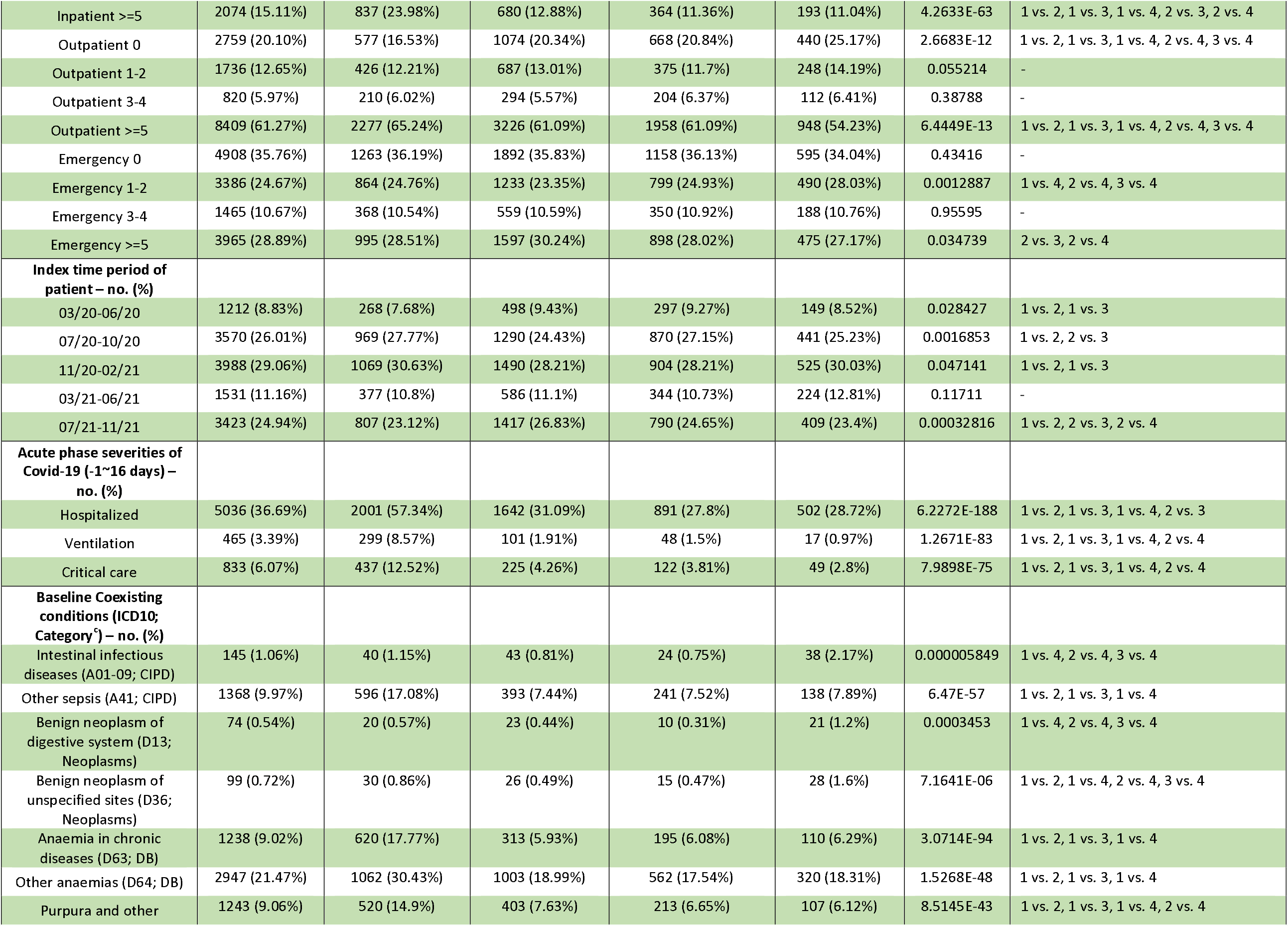
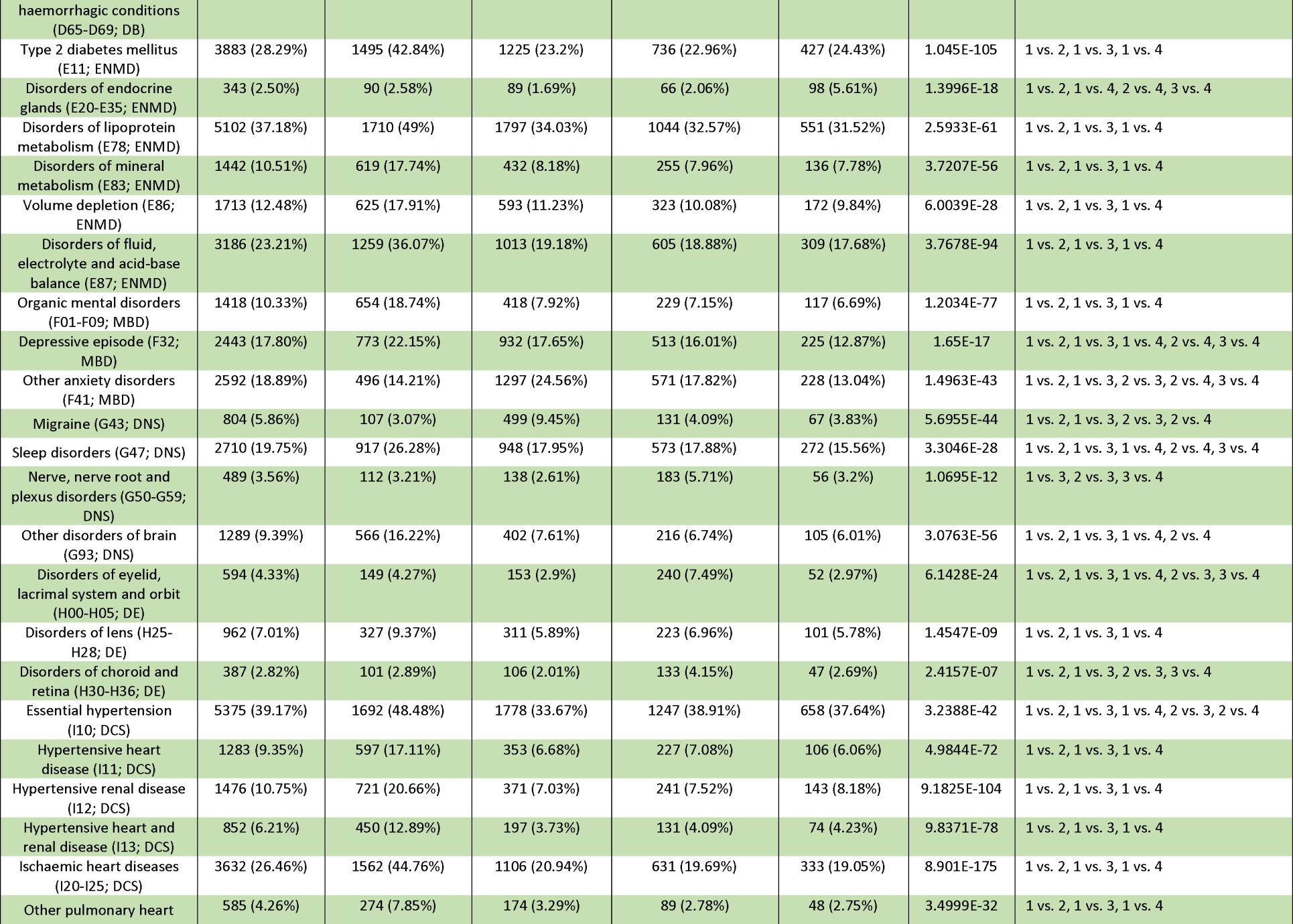
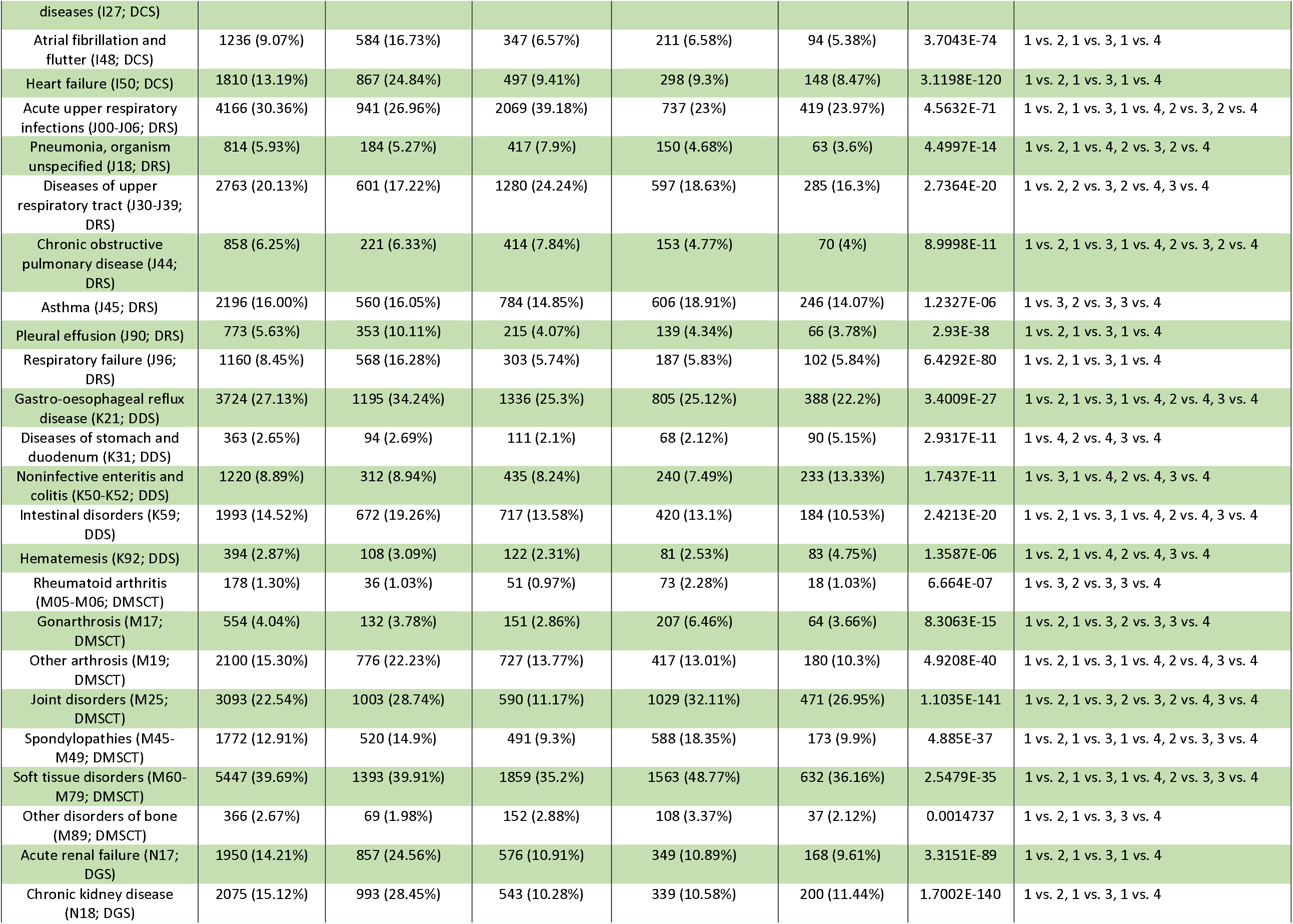
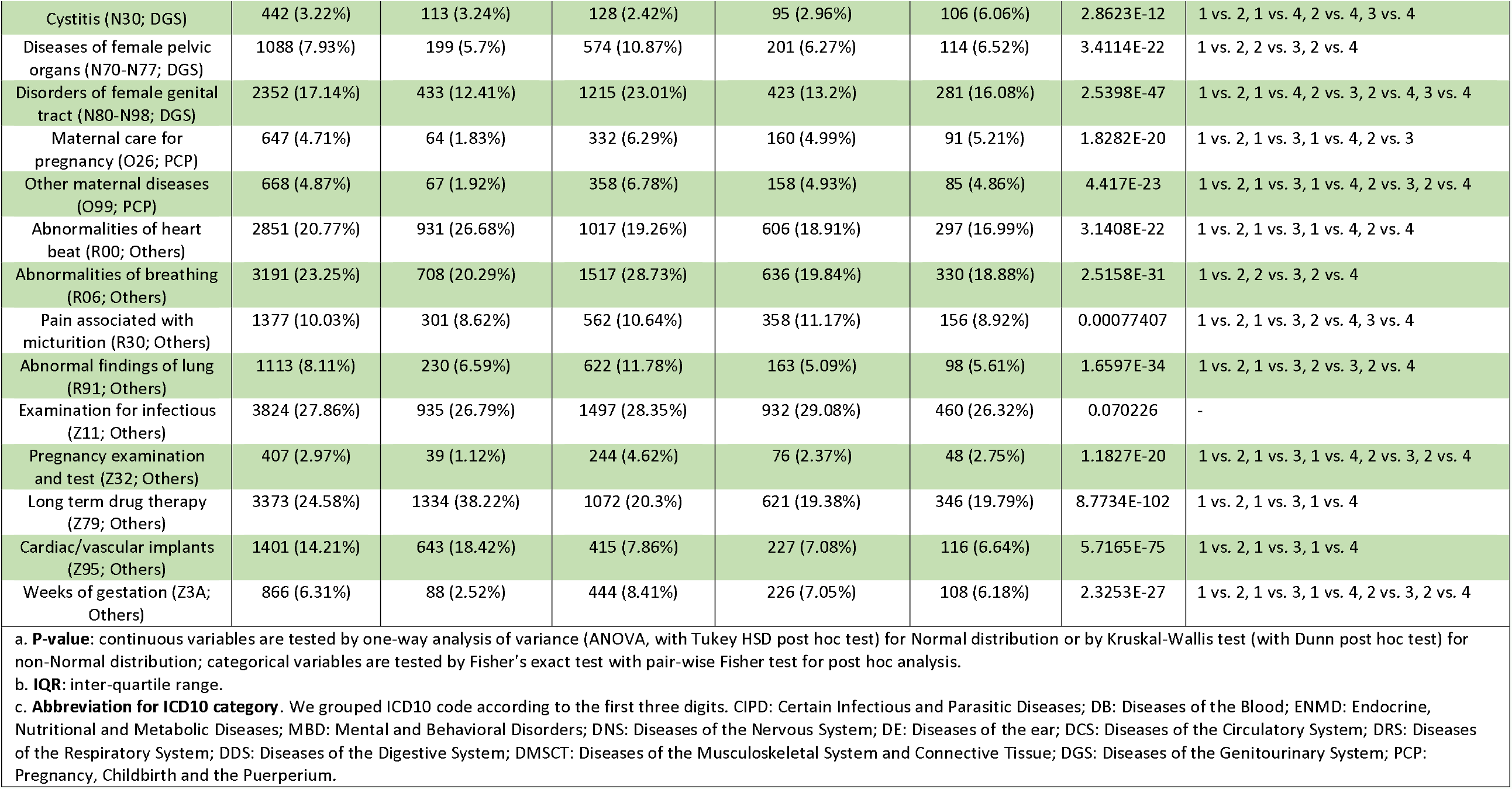
Characteristics of the identified subphenotypes on the OneFlorida+ cohort.

**Supplemental Figure 1.**
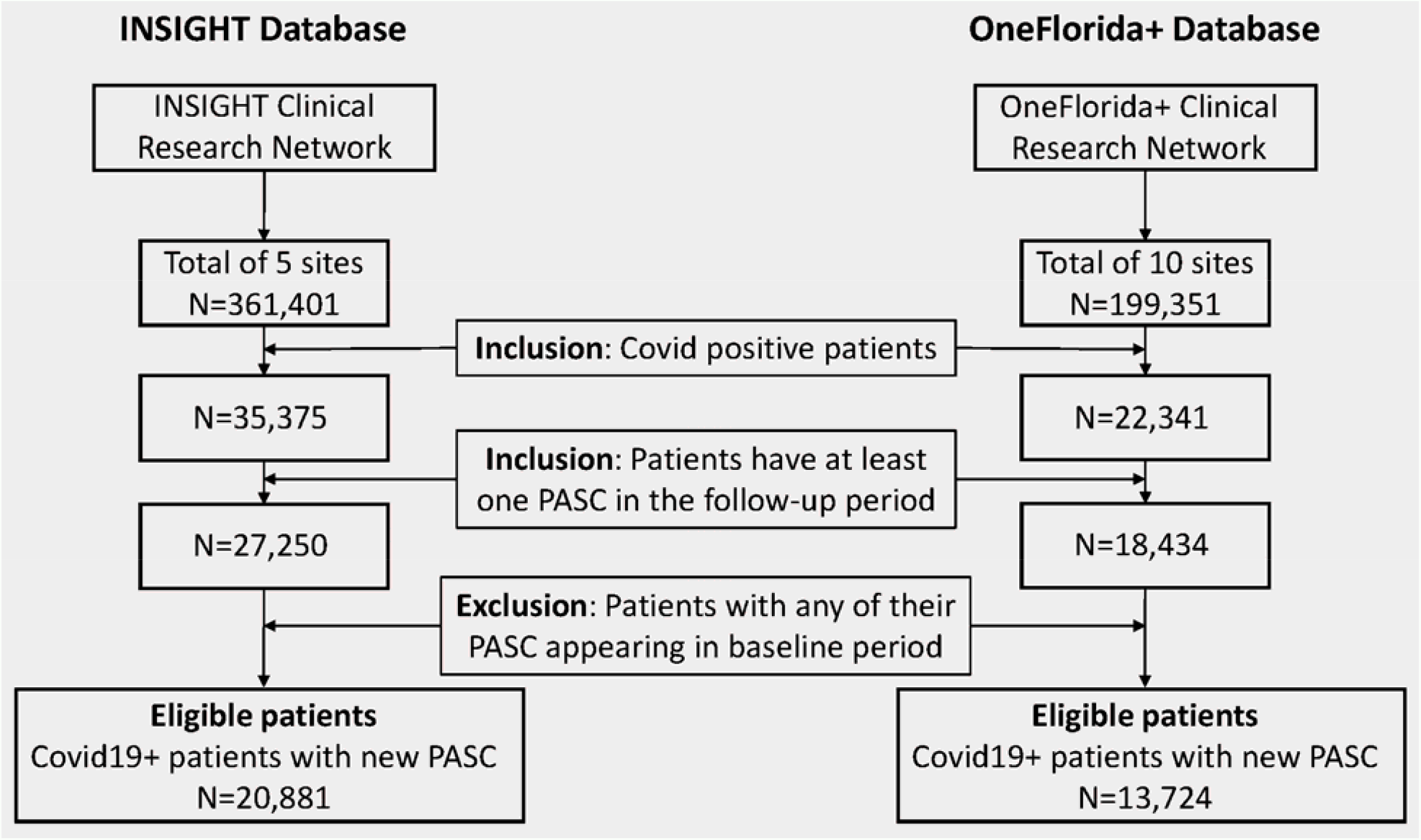
Inclusion-exclusion cascade for the INSIGHT and OneFlorida+ cohorts.

**Supplemental Figure 2.**
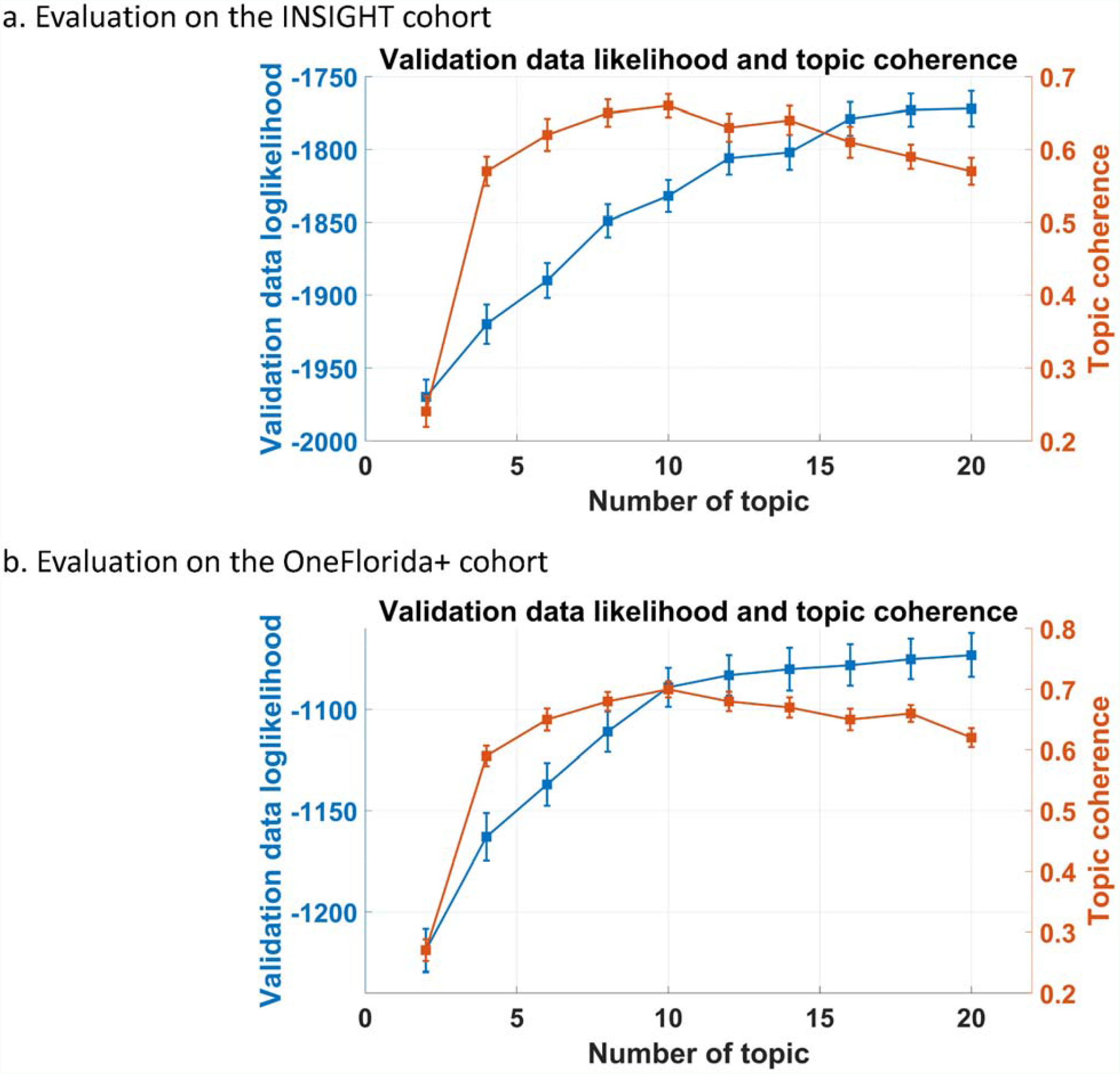
The data likelihood and topic coherence conditioned on different number of topics, which were regarded as the criteria to select the optimal number of topics.

**Supplemental Figure 3.**
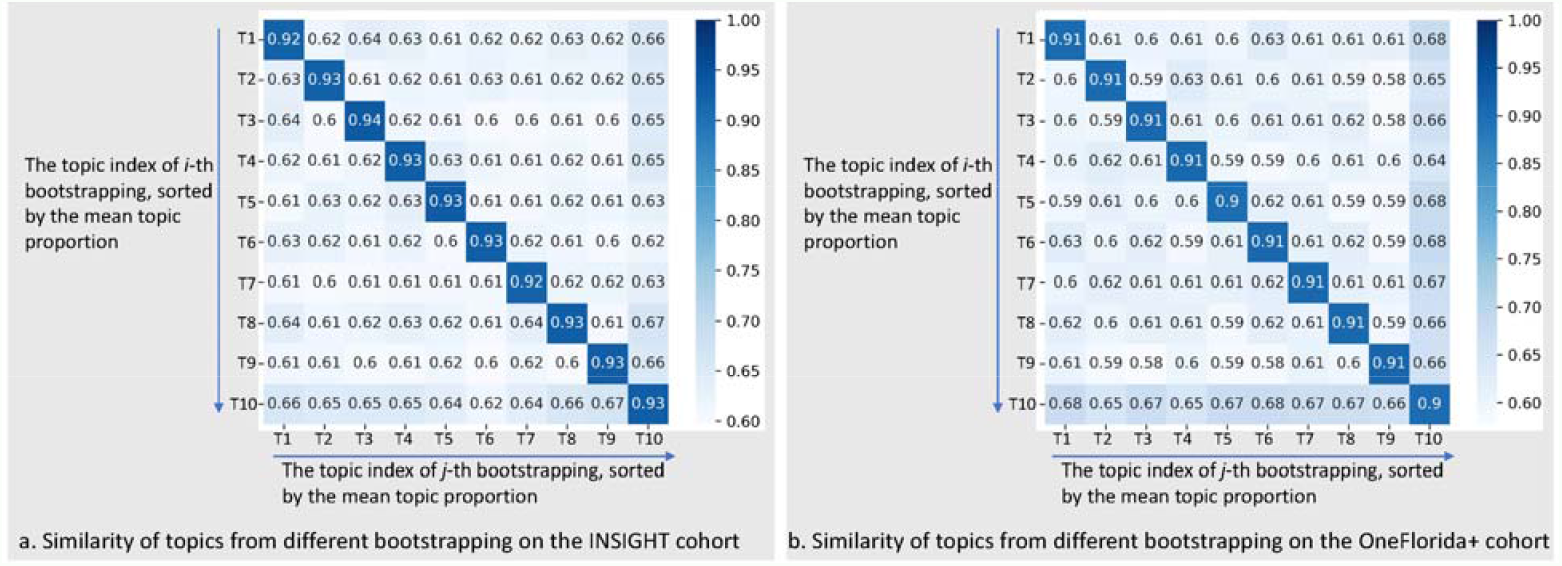
The similarity of topics from different bootstrapping, which were used to evaluate the robustness of topics.

**Supplemental Figure 4.**
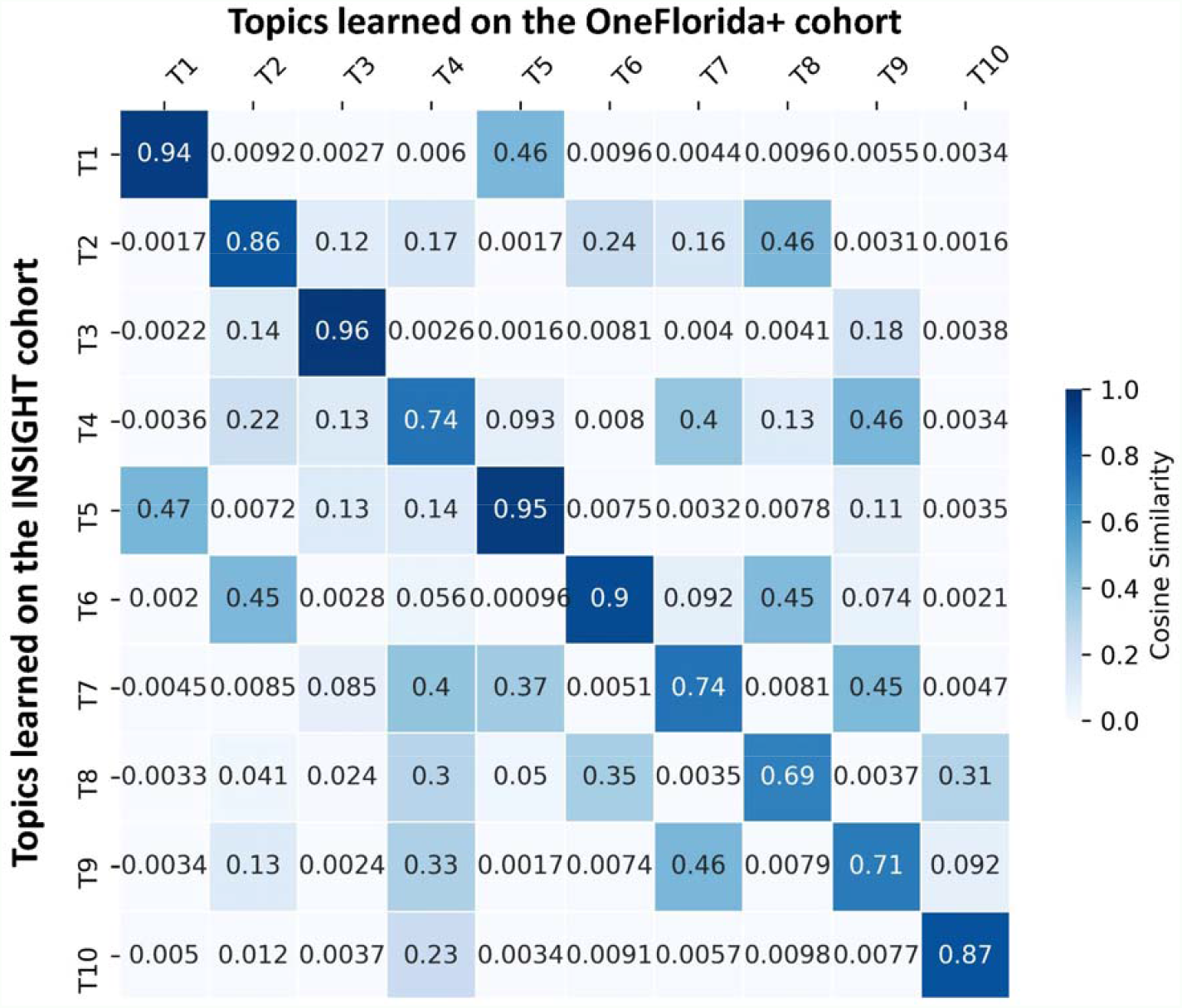
The similarity of topics from two cohorts, which were used to evaluate the overlap of topics learned on two cohorts.

**Supplemental Figure 5.**
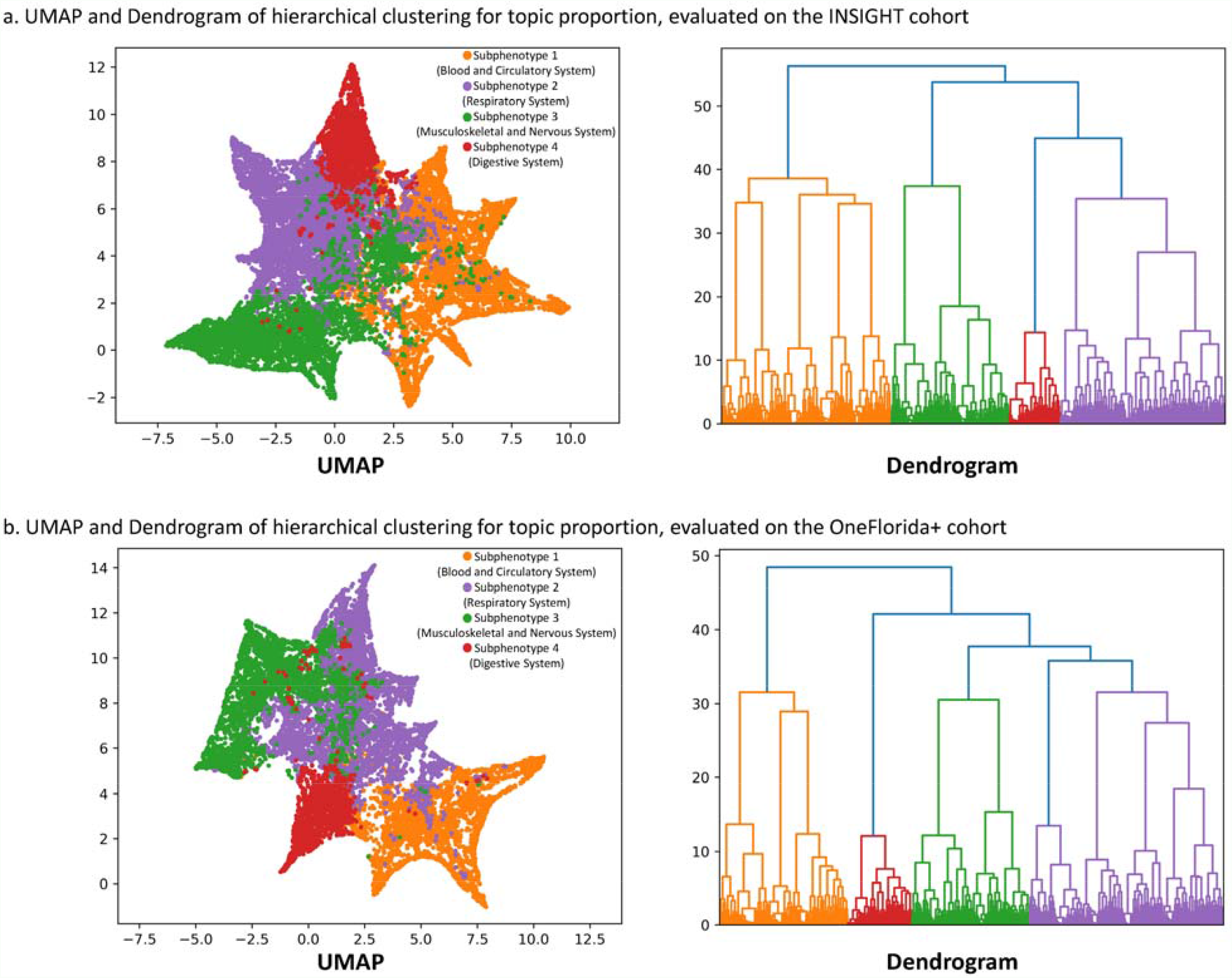
UMAP and dendrogram for two cohorts.

**Supplemental Figure 6.**
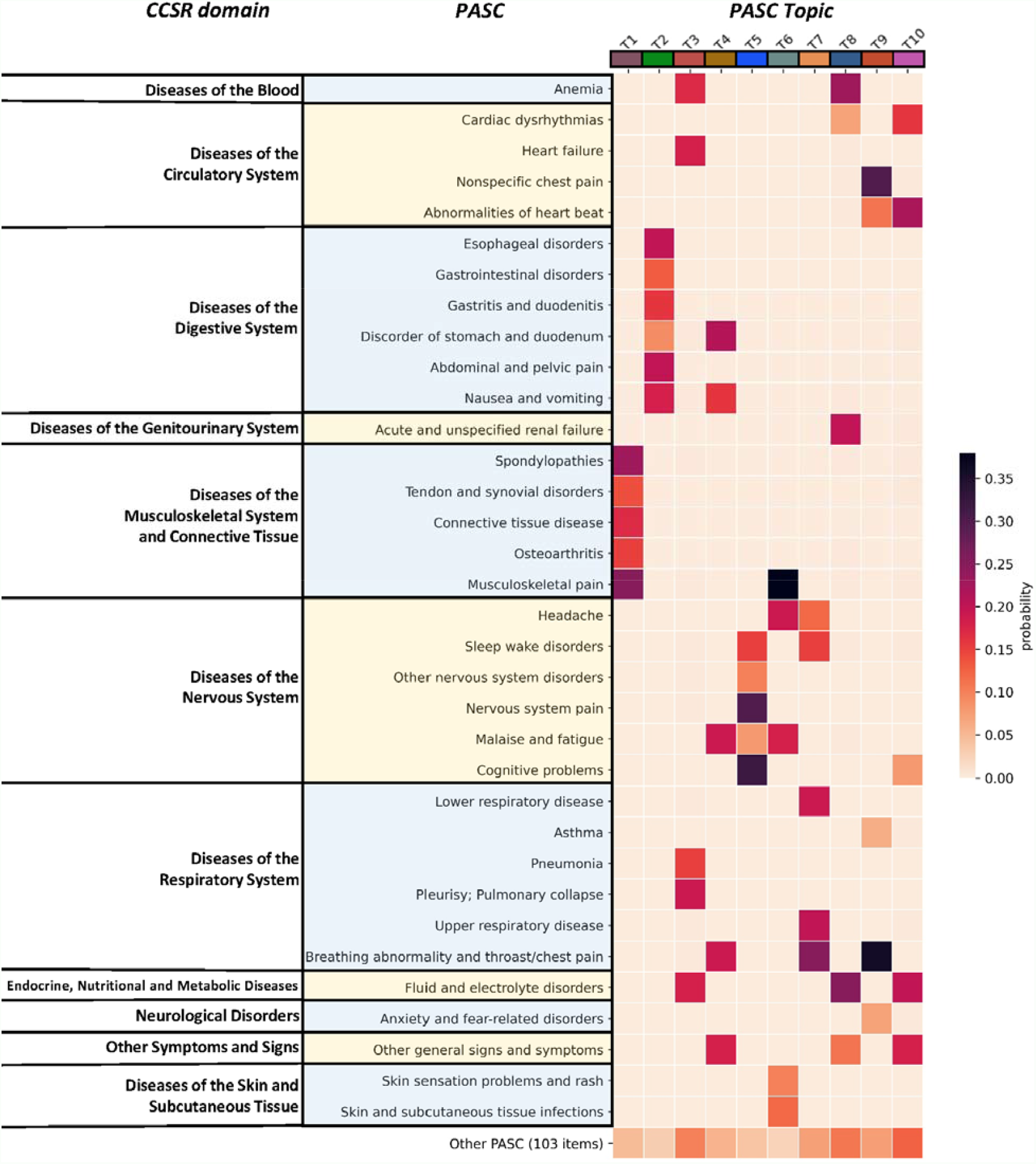
The heatmap of PASC topics learned on the OneFlorida+ cohort. Each row denotes a potential PASC category grouped by different CCSR domains, and each column denotes a particular PASC topic. Each PASC topic is characterized by a unique post-acute incidence probability distribution over all 137 individual potential PASC categories.

**Supplemental Figure 7.**
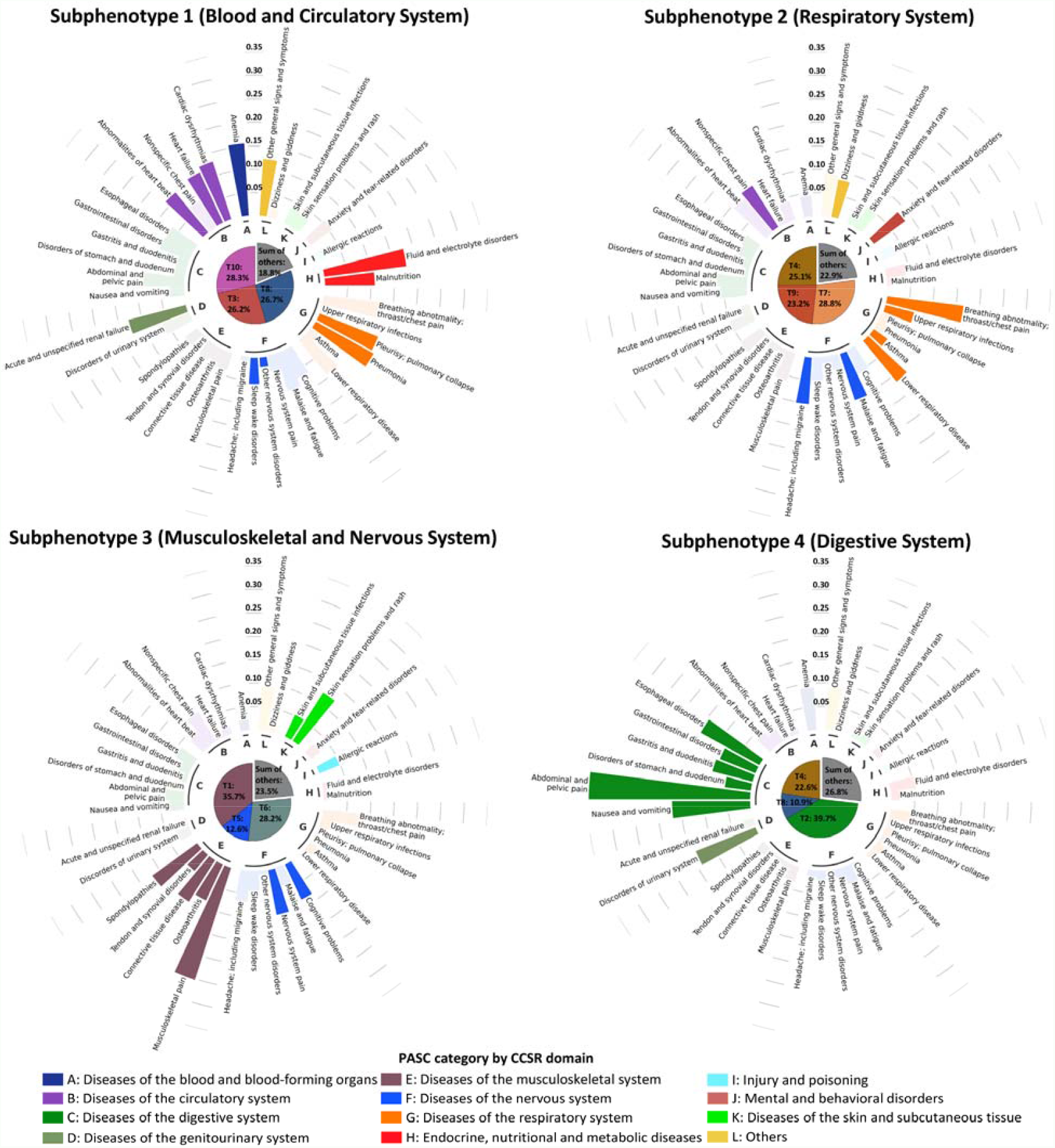
The prevalence (denoted by the percentage of patients in this subphenotype having each PASC condition) of PASC conditions for each subphenotype on the OneFlorida+ cohort, where PASC conditions were grouped into different categories shown by different colors. For one PASC condition, if it is most prevalent in one subphenotype, we highlighted it in this subphenotype. In the center of each plot, we used pie chart to represent the mean topic proportions on this subphenotype.

**Supplemental Figure 8.**
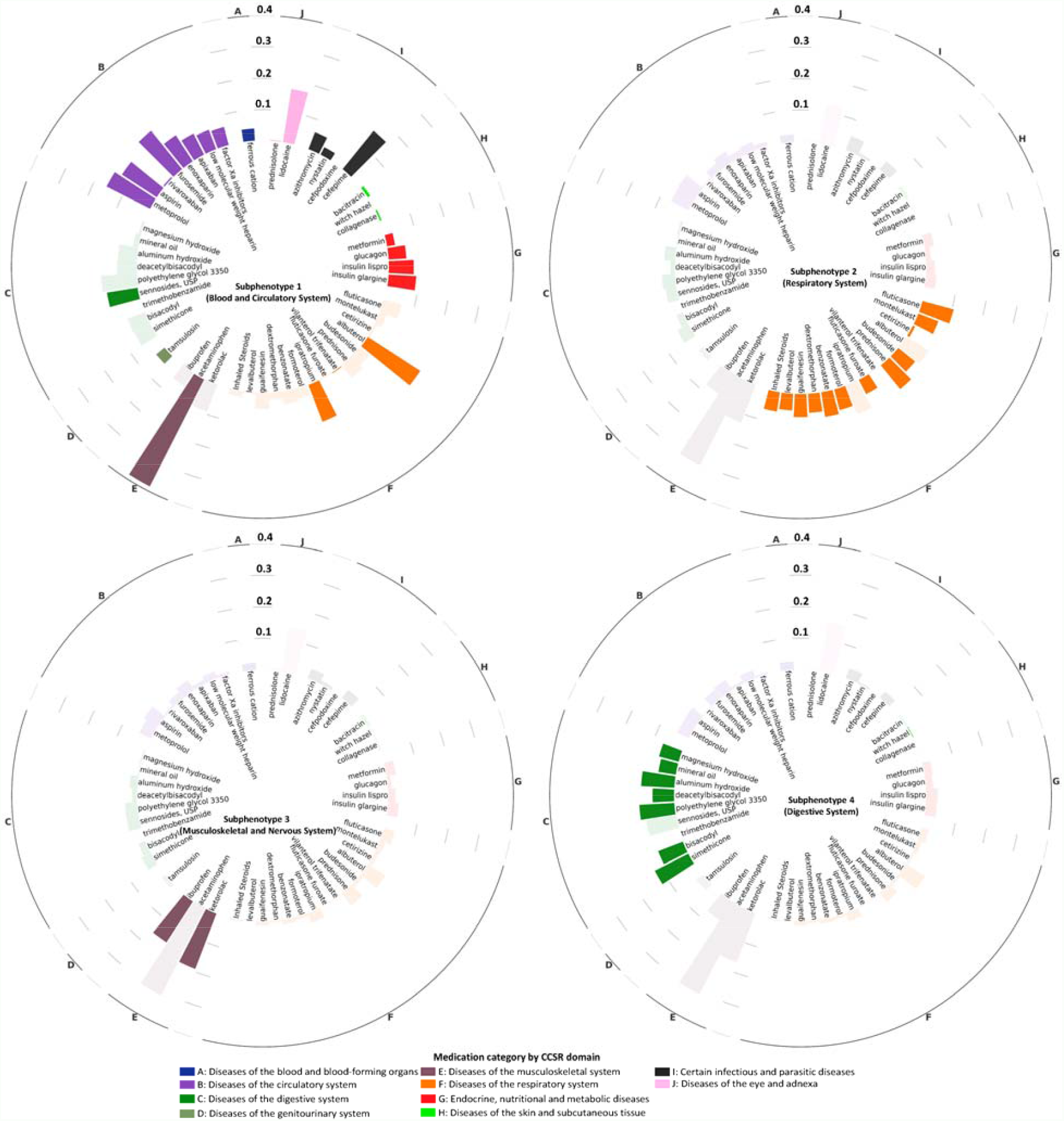
The prevalence of incident prescriptions of medications in the post-acute infection period for each subphenotype on the OneFlorida+ cohort, where medications are grouped into different categories shown by different colors. For one of the medications, if it is most prevalent in one subphenotype, we highlighted it in this subphenotype.

**Supplemental Figure 9.**
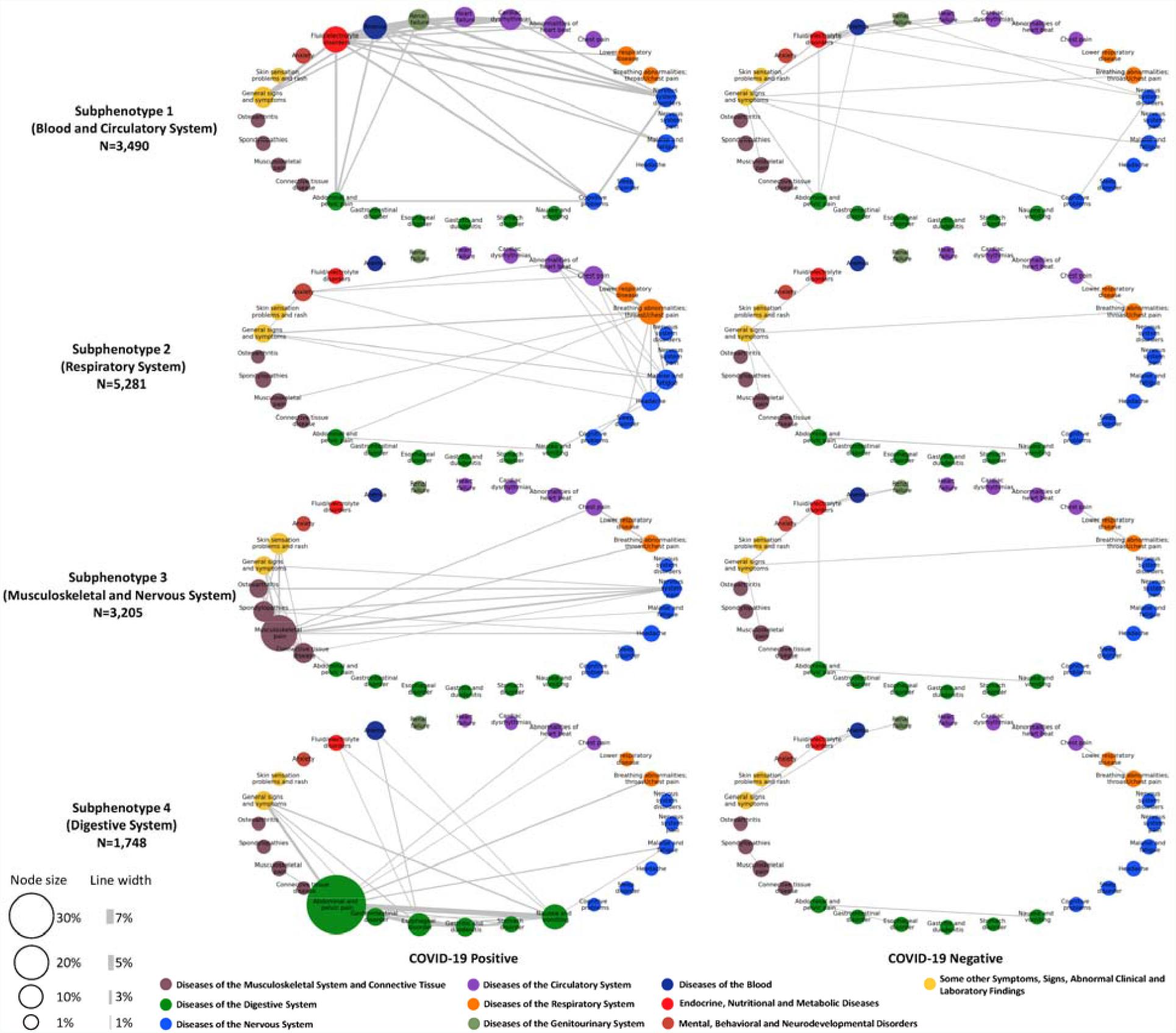
Difference of the incidence patterns of selected PASC conditions (grouped by CCSR domains) in 30-180 days after COVID-19 lab test between positive and matched negative patients on the OneFlorida+ cohort. The bubbles in each network correspond to a PASC condition with their sizes proportional to the incidence rate in the particular subphenotype or matched controls. The edge linking a pair of bubbles indicates co-incidence of the corresponding investigative conditions with its thickness proportional to the co-incidence rate, where we showed the line if the rate is larger than 1%.

## Supplemental Methods

### Evaluate the data likelihood and topic coherence

To select the optimal number of topics, based on different number of topics, we learned topic model and calculated the data likelihood and topic coherence (Supplemental Figure 2) according to the following methods.

### Data likelihood

In PFA, to model the binary PASC vector, we used the Bernoulli-Poisson link as:

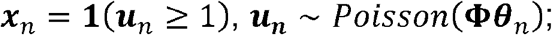

where,***x***_*n*_ is the binary PASC vector, **Φ** is the topic matrix, ***θ***_*n*_ is the topic proportion vector, ***u***_*n*_ is the latent variables which links the binary observation and topic representation. According to the property of Bernoulli-Poisson link, ***u***_*n*_ can be marginalized out and then one can obtain a Bernoulli likelihood as

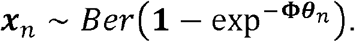

Given 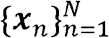, after learning **Φ** and 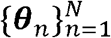, we can calculate the Bernoulli data likelihood.

### Topic coherence

Topic coherence is an important matric to evaluate the quality of topics based on the input data. It is measured based on a sliding window (in our case, the length of the window is the total number of PASCs), and then one can calculate the normalized pointwise mutual information (NPMI) between input data and the learned topics. We used the python package GENSIM (https://radimrehurek.com/gensim/) to calculate the topic coherence.

